# Early use of nitazoxanide in mild Covid-19 disease: randomized, placebo-controlled trial

**DOI:** 10.1101/2020.10.21.20217208

**Authors:** Patricia R. M. Rocco, Pedro L. Silva, Fernanda F. Cruz, Marco Antonio C. M. Junior, Paulo F. G. M. M. Tierno, Marcos A. Moura, Luís Frederico G. De Oliveira, Cristiano C. Lima, Ezequiel A. Dos Santos, Walter F. Junior, Ana Paula S. M. Fernandes, Kleber G. Franchini, Erick Magri, Nara F. de Moraes, José Mário J. Gonçalves, Melanie N. Carbonieri, Ivonise S. Dos Santos, Natália F. Paes, Paula V. M. Maciel, Raissa P. Rocha, Alex F. de Carvalho, Pedro Augusto Alves, José Luiz P. Modena, Artur T. Cordeiro, Daniela B. B. Trivella, Rafael E. Marques, Ronir R. Luiz, Paolo Pelosi, Jose Roberto Lapa e Silva, on behalf of the SARITA-2 investigators

## Abstract

The antiparasitic drug nitazoxanide is widely available and exerts broad-spectrum antiviral activity *in vitro*. However, there is no evidence of its impact on SARS-CoV-2 infection.

In a multicenter, randomized, double-blind, placebo-controlled trial, adult patients who presented up to 3 days after onset of Covid-19 symptoms (dry cough, fever, and/or fatigue) were enrolled. After confirmation of SARS-CoV2 infection by RT-PCR on nasopharyngeal swab, patients were randomized 1:1 to receive either nitazoxanide (500 mg) or placebo, TID, for 5 days. The primary outcome was complete resolution of symptoms. Secondary outcomes were viral load, general laboratory tests, serum biomarkers of inflammation, and hospitalization rate. Adverse events were also assessed.

From June 8 to August 20, 2020, 1,575 patients were screened. Of these, 392 (198 placebo, 194 nitazoxanide) were analyzed. Median time from symptom onset to first dose of study drug was 5 (4-5) days. At the 5-day study visit, symptom resolution did not differ between the nitazoxanide and placebo arms. However, at the 1-week follow-up, 78% in the nitazoxanide arm and 57% in the placebo arm reported complete resolution of symptoms (*p*=0.048). Swabs collected were negative for SARS-CoV-2 in 29.9% of patients in the nitazoxanide arm *versus* 18.2% in the placebo arm (p=0.009). Viral load was also reduced after nitazoxanide compared to placebo (p=0.006). No serious adverse events were observed.

In patients with mild Covid-19, symptom resolution did not differ between the nitazoxanide and placebo groups after 5 days of therapy. However, early nitazoxanide therapy was safe and reduced viral load significantly.

**Take home message:** This was the first study to evaluate the effect of early nitazoxanide therapy in mild Covid-19. Nitazoxanide did not accelerate symptom resolution after 5 days of therapy; however, reduced viral load significantly with no serious adverse events.

## Introduction

The majority of patients infected with the severe acute respiratory syndrome coronavirus-2 (SARS-CoV-2) present mild symptoms of coronavirus disease 2019 (Covid-19) and recover with supportive care; however, in certain hosts, even mild disease may progress to clinical deterioration or a protracted course [1]. To date, no therapeutic interventions have proven effective in mild Covid-19.

Drug repurposing has been recognized as an important tool in the search for effective Covid-19 therapies, as it reduces development costs and timelines [2]. Nitazoxanide, a clinically approved and commercially available antiparasitic drug, has been found to have broad-spectrum antiviral activity, including against coronaviruses, influenza viruses, and hepatitis B and C viruses [3]. Furthermore, it has been shown to inhibit SARS-CoV-2 replication at low micromolar concentrations in Vero CCL81 cells [4]. In addition, nitazoxanide is orally bioavailable and broadly well-tolerated, thus representing a promising alternative for the management of Covid-19 were it to prove effective *in vivo* [3]. Efficacy in early treatment—initiated soon after symptom onset to reduce viral load and prevent disease progression—would be particularly appealing from a public health standpoint. However, there is no evidence of its safety or efficacy as therapy for mild Covid-19 patients.

In this context, a multicenter, randomized, placebo-controlled trial was carried out to evaluate whether early nitazoxanide therapy would be effective in accelerating symptom resolution. Secondarily, viral load, markers of inflammation, hospitalization rate, and the safety of nitazoxanide as compared with placebo were also assessed.

## Methods

### Study design

A double-blind, placebo-controlled trial was conducted at 5 freestanding urgent care centers and 2 hospitals across Brazil (**Supplementary Table S1**). The trial was designed by the executive committee and approved by the Brazilian National Commission for Research Ethics and individual ethics committees of the participating sites (CAAE: 32258920.0.1001.5257). The trial was funded by the Brazilian Ministry of Science, Technology, and Innovation via the National Council for Scientific and Technological Development (CNPq). Nitazoxanide was provided free of charge by Eurofarma, which had no further role in the design or conduct of the trial. The executive committee assures the accuracy of the data and fidelity of the trial to the protocol, which was registered in the Brazilian Registry of Clinical Trials (REBEC) number RBR-4nr86m and ClinicalTrials.gov number NCT04552483. The independent Data and Safety Monitoring Board (DSMB), composed of experts in clinical trials and infectious diseases, was convened after 25%, 50%, and 75% of the participants had completed 14 days of follow-up and had access to information on adverse events and efficacy outcomes at every quartile.

The trial was conducted in accordance with the principles of the Declaration of Helsinki and the International Conference on Harmonization (ICH) Good Clinical Practice Guideline (E6R2). Online clinical monitoring and quality control were outsourced to a contract research organization (ATCGen, Campinas, Brazil). This report follows the Consolidated Standards of Reporting Trials (CONSORT) guideline [5]. The final protocol, amendments and changes to the trial protocol, and the final statistical analysis plan are detailed in the Supplemental Methods.

### Patients

Consecutive adult patients (aged 18 years or older) who presented with clinical symptoms of Covid-19 (defined for the purposes of this trial as dry cough, fever, and/or fatigue) of no longer than 3 days’ duration were enrolled. The exclusion criteria were: negative reverse-transcriptase quantitative real-time polymerase chain reaction (RT-PCR) test for SARS-CoV-2 on an nasopharyngeal swab specimen; inability to swallow; preexisting conditions precluding the safe conduct of study procedures, including severe renal, heart, respiratory, liver, or autoimmune diseases, cancer in the last 5 years, or known allergy or hypersensitivity to nitazoxanide; therapy with nitazoxanide in the 30 days before presentation; and clinical suspicion of bacterial pneumonia or tuberculosis.

### Randomization and masking

Patients were randomly assigned (1:1) using a computer-generated random number list to receive either placebo or nitazoxanide (500 mg oral solution, 20 mg/mL [25 mL], three times daily for 5 days), dispensed by the pharmacy of each study site. Placebo and nitazoxanide were color-matched to ensure that assessors were unaware of group allocation at all time points.

### Procedures

On day 1 (baseline), patients were assessed for eligibility. Informed consent was obtained from each patient or, if the patient was unable to provide consent, from a healthcare proxy. A nasopharyngeal swab was then collected for RT-PCR testing. In order to mitigate any bias, all RT-PCR analyses were processed centrally at CT-VACINAS, Federal University of Minas Gerais, Brazil; specimens were sent on the day of collection by commercial courier. Patients who tested positive for SARS-CoV-2 (result obtained 1-2 days after RT-PCR) were contacted by telephone and asked to return to the health facility to which they had originally presented. Site investigators then performed a comprehensive physical examination and measured body temperature, heart and respiratory rates, blood pressure, and peripheral oxygen saturation. Ethnicity, current medications, and date of symptom onset were self-reported by patients. Blood was then drawn for measurement of complete blood count, C-reactive protein (CRP), and serum biomarkers of inflammation (interleukin [IL]-6, IL-8, IL-1β, tumor necrosis factor [TNF]-α, and interferon [IFN]-γ) **(See Supplemental Methods for details of RT-PCR testing and biomarker assessment)**.

All patients took home a symptom journal designed to gather information on daily symptoms, new symptoms, and the date of resolution of each symptom (**Supplemental Table S2**). Study data were entered directly into an electronic database by an assigned staff member at each study site and further validated by external trial monitoring staff at ATCGen.

One day after completion of therapy, patients returned to the study sites to return their symptom journals and provide a new nasopharyngeal sample for RT-PCR and blood samples for complete blood count, CRP, and serum biomarkers. Patients who remained symptomatic at the end-of-therapy visit were contacted by telephone 1 week later for further assessment. Data validation were done exclusively by the designated study site manager, with assistance from ATCGen staff as needed. If any delay was detected in nasopharyngeal swab sample analysis (i.e., if results were unavailable on day 3), the study protocol accepted extension of treatment initiation on day 4, but no later.

### Primary Outcome

The primary outcome was complete resolution of the three symptoms of interest (dry cough, fever, and/or fatigue) after therapy.

### Secondary Outcomes

Secondary outcomes were reduction in viral RNA load on nasopharyngeal swab specimens (from baseline until the day after completion of therapy); improvement in laboratory parameters (including serum biomarkers of inflammation); and incidence of hospital admission after completion of therapy. Patients who did not return to the study sites after the end of therapy were contacted by telephone to understand the reasons for nonadherence and were then excluded from per-protocol analysis (**Supplemental Methods**). Adverse events, regardless of grade, were monitored throughout the trial by review of the electronic medical record, physical examination, vital signs, and laboratory tests from enrollment through day 14. The Medical Dictionary for Regulatory Activities® (MedDRA, version 23.0) was used for classification.

### Statistical Analyses

We estimated a sample size of 392 patients (196 per arm, allocation ratio 1:1) would provide 85% power to detect an 11% increase in symptom-free days after nitazoxanide initiation compared to placebo at a two-sided significance level of α=0.05. Descriptive statistics were used for demographic, laboratory, and clinical data. For qualitative variables, Fisher’s exact test was performed. The Mann-Whitney *U* and Wilcoxon tests were used for between and within-groups comparisons, respectively. Statistical analyses were performed in the SPSS Version 27 environment (IBM Corp, Armonk, NY), and a two-tailed *p*-value < 0.05 was considered significant **(Supplemental Methods)**.

## Results

Between June 8 and August 20, 2020, 1,575 patients were assessed for eligibility at the study sites. Of these, 475 tested positive for SARS-CoV-2 infection by RT-PCR and underwent randomization. **Figure 1** summarizes the enrollment and follow-up of study participants. Reasons for exclusion before randomization included negative RT-PCR collected at day 1 and/or absence of Covid-19 symptoms (n=1,062), refusal to participate (n=27), hospitalization before the first dose of therapy (n=5), and other reasons (n=6). After randomization (n=475), patients were excluded due to discontinued intervention (n=41), adverse events (n=7), and hospitalization (n=10). During analysis, 12 patients were excluded from the nitazoxanide arm and 15 from the placebo arm due to protocol deviation, missing data on the primary outcome, or non-evaluability, resulting in a studied population of 392 patients (194 in the nitazoxanide arm and 198 in the placebo arm).

**Figure 1.**
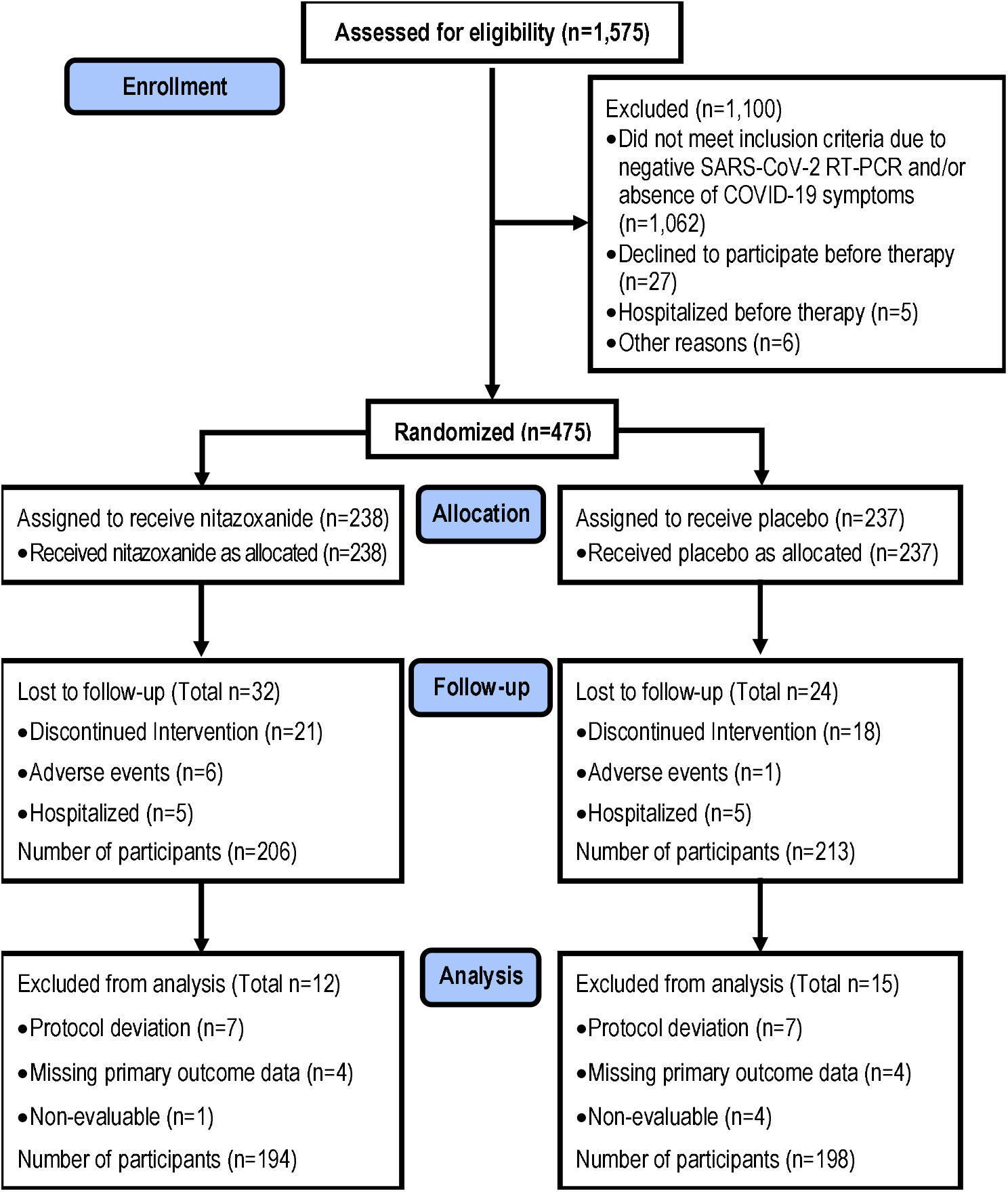
Enrollment, Randomization, Follow-up, and Treatment. 1,575 patients were assessed for eligibility at the study sites. Of these, 475 tested positive for SARS-CoV-2 infection by RT-PCR and underwent randomization. Reasons for exclusion before randomization included negative RT-PCR collected at day 1 (baseline) and/or absence of Covid-19 symptoms (n=1,062), refusal to participate (n=27) and hospitalization before the first dose of therapy (n=5), and other reasons (n=6). After randomization (n=475), patients were excluded due to discontinued intervention (n=39), moderate adverse events (n=7), and hospitalization (n=10). During analysis, 12 patients were excluded from the nitazoxanide arm and 15 from the placebo arm due to protocol deviation, missing data on the primary outcome, or non-evaluability, resulting in a studied population of 392 patients (194 in the nitazoxanide arm and 198 in the placebo arm).

### Baseline characteristics

Patients’ characteristics at baseline are given in **Table 1**. Demographics and clinical characteristics were balanced in both groups. Age ranged from 18 to 77 years. Just over half (53%) were women and 69% were white. Regarding timing of therapy initiation, 8% of patients were enrolled on day 1, 25% on day 2, and 67% on day 3 after symptom onset. The median (IQR) time between symptom onset and first dose of nitazoxanide or placebo was 5 (4-5) days. Fever, dry cough, and fatigue were the most frequent clinical features present at enrollment. At baseline, the overall median (IQR) viral load in the nasopharyngeal swab at baseline did not differ significantly between the two arms Additional characteristics of the study population at baseline are given in **Table S3**.

**Table 1.**
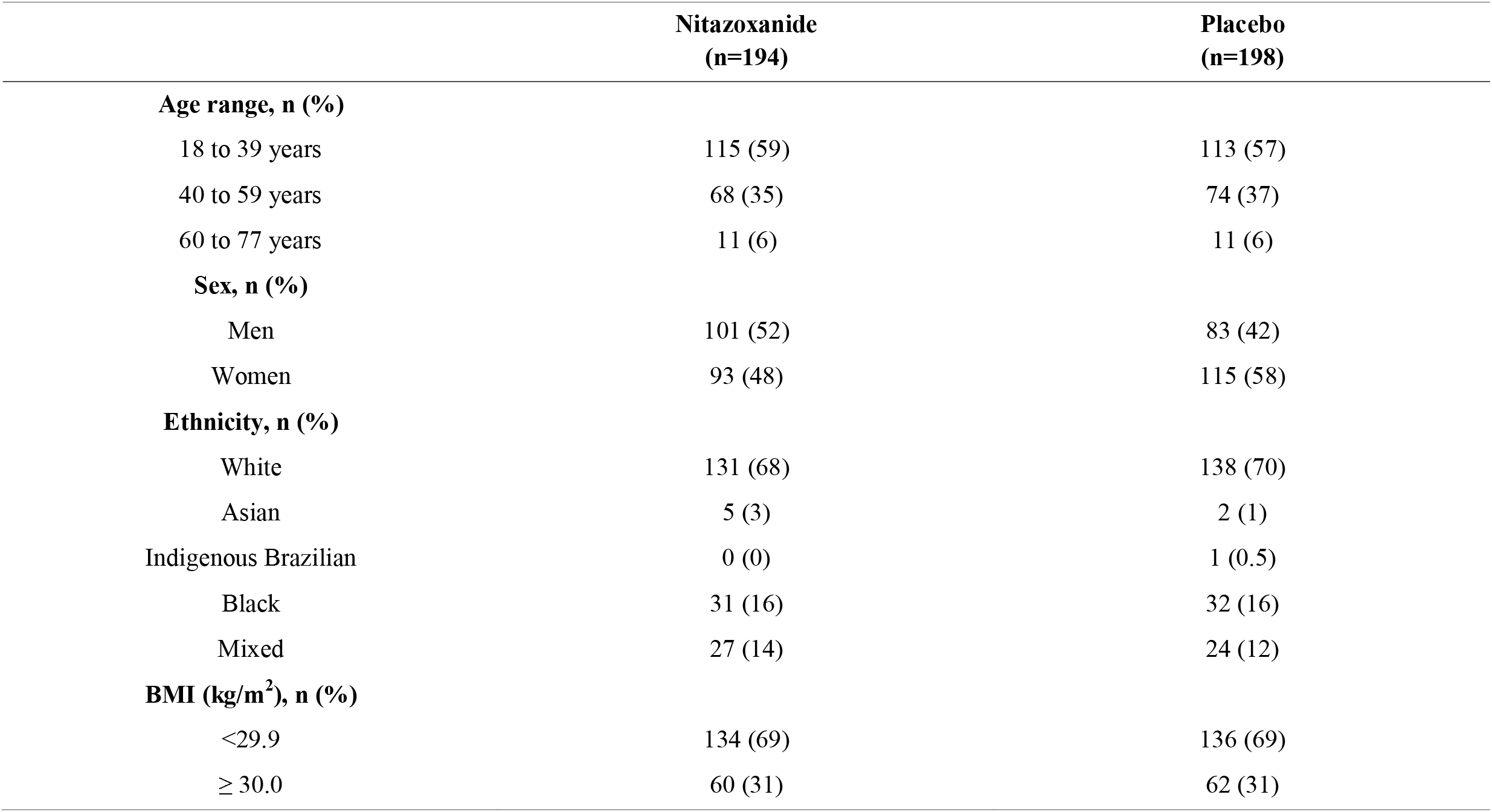
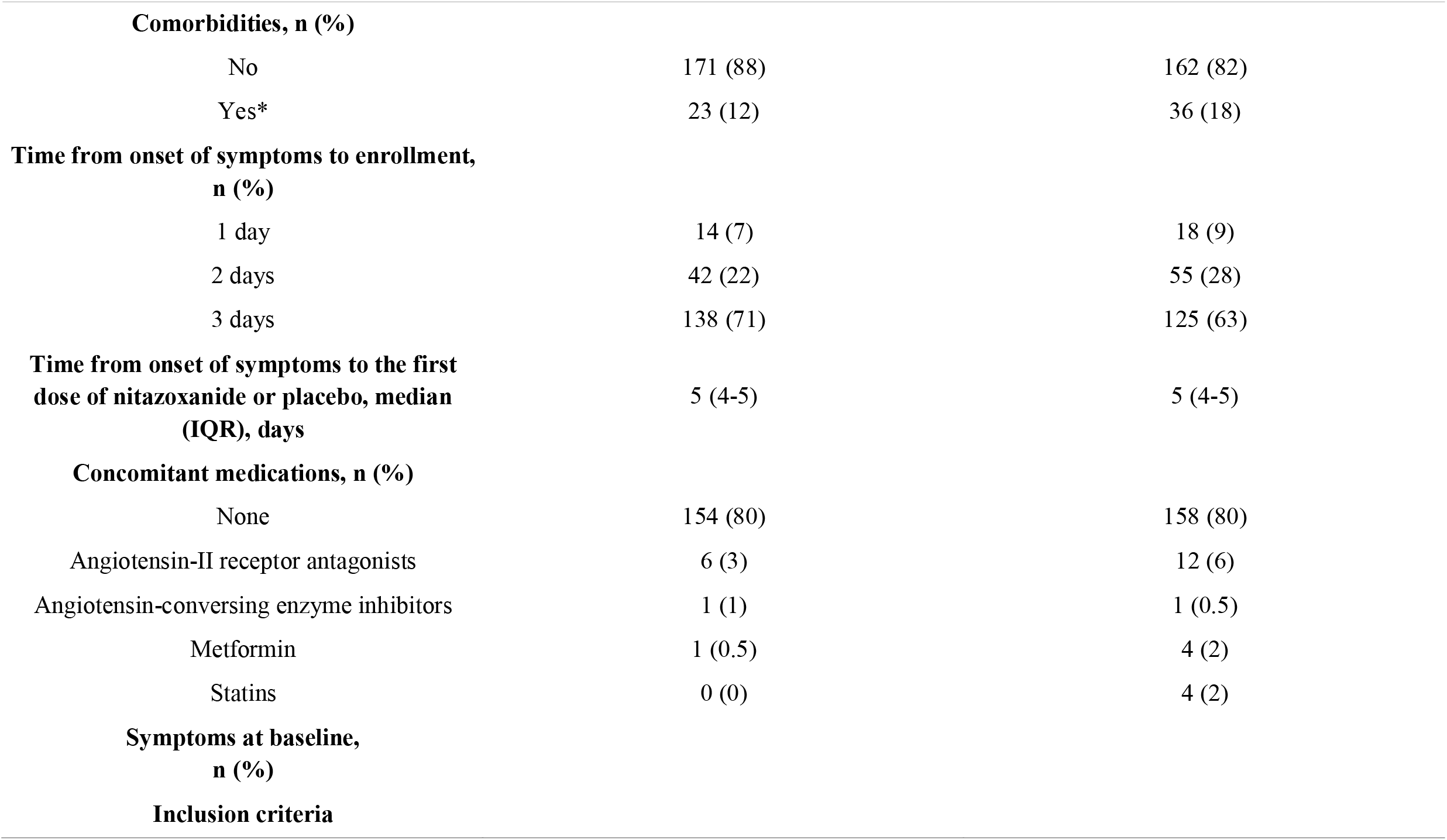
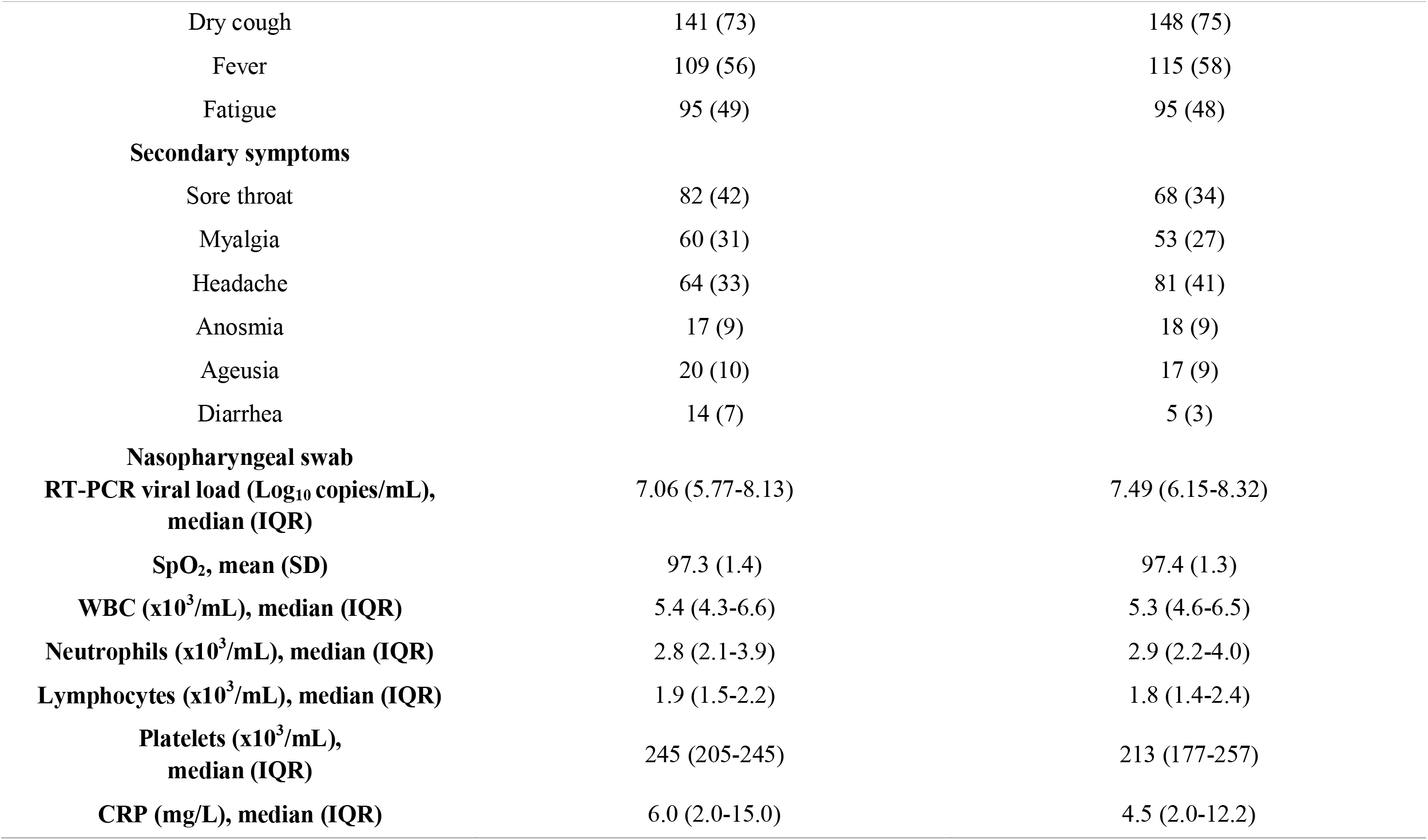
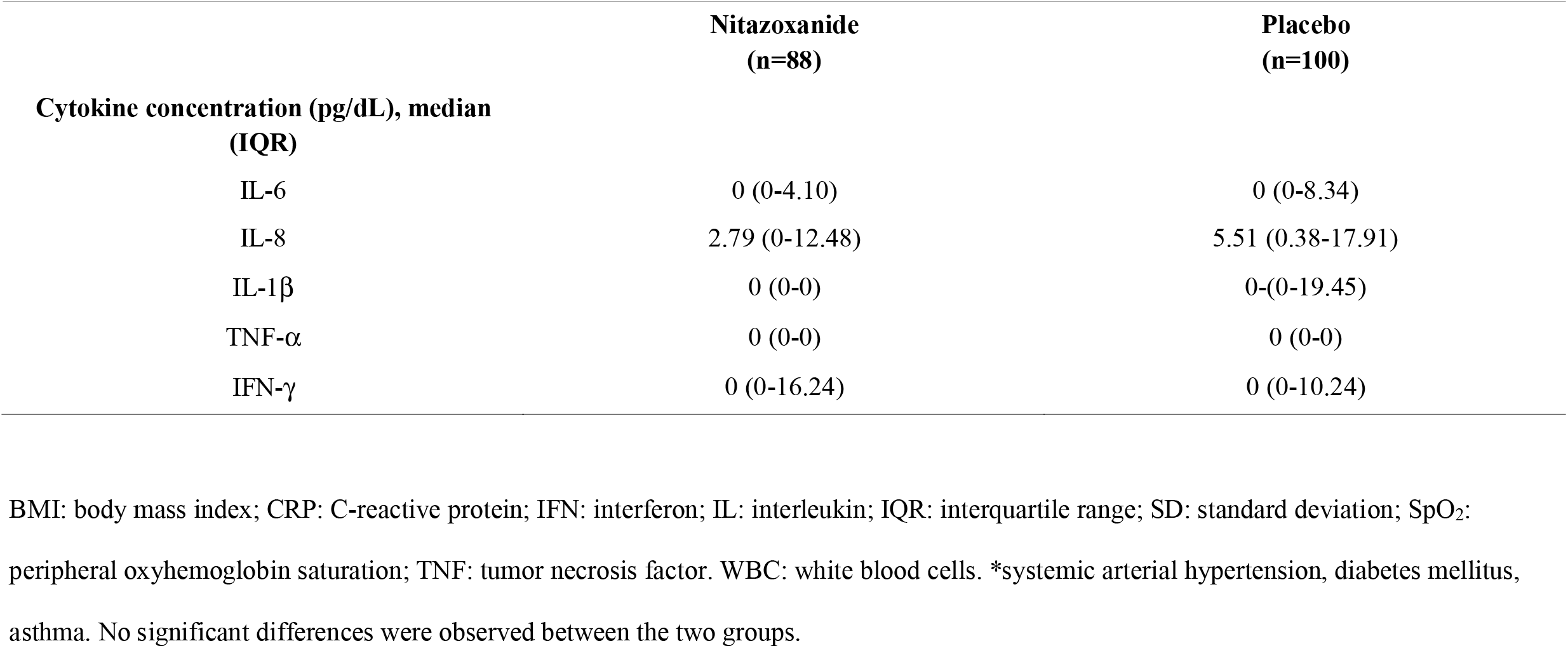
Characteristics of the Participants Testing Positive for SARS-CoV-2 on RT-PCR at Baseline.

### Primary Outcome

Complete resolution of symptoms (dry cough, fever, and fatigue) did not differ between the nitazoxanide and placebo arms after 5 days of therapy **(Table 2, Figure S2)**. Overall, 49 patients in the nitazoxanide arm and 46 in the placebo arm continued to report at least one of the three symptoms of interest on completion of therapy. At the 1-week follow-up phone call, 38 patients (78%) in the nitazoxanide arm and 26 (57%) in the placebo arm reported complete resolution of symptoms (*p*=0.048).

**Table 2.** Primary outcome (free from all symptoms) and components (dry cough, fever, and fatigue) after 5 days of therapy.

|  | **Nitazoxanide**<br>**(n=194)** | **Placebo**<br>**(n=198)** | ***p value*** |
| --- | --- | --- | --- |
| **Primary outcome** |  |  |  |
| **All symptoms** | 135 (70) | 146 (74) | 0.423 |
| **Components of primary outcome** |  |  |  |
| **Dry cough** | 110 (78) | 119 (80) | 0.721 |
| **Fever** | 97 (89) | 110 (96) | 0.101 |
| **Fatigue** | 67 (71) | 69 (73) | 0.872 |

Data are n (%).

### Secondary Outcome

After 5 days of therapy, viral load was lower in the nitazoxanide arm compared to placebo (*p*=0.006) **(Table 3)**. Moreover, the percentage of reduction in viral load from day 1 (baseline) **(Table 1)** to day 5 **(Table 3)** of therapy was significantly higher in the nitazoxanide arm (55%) than in the placebo arm (45%) (*p*=0.013). At the end-of-therapy visit, 29.9% of patients in the nitazoxanide arm *versus* 18.2% in the placebo arm were negative for SARS-CoV-2 on RT-PCR (*p*=0.009) **(Table 3)**. Vital signs **(Table S3)**, total leukocyte, neutrophil, lymphocyte, and platelet counts, CRP levels, and serum biomarkers of inflammation **(Table 3)** did not differ between baseline and day 5 of therapy in the nitazoxanide arm, nor between the nitazoxanide and placebo arms. Ten patients (5 from each arm) were hospitalized due to clinical deterioration; none had completed the 5-day course of therapy. Two patients, both in the nitazoxanide arm, required intensive care unit admission **(Table S4)**.

**Table 3.**
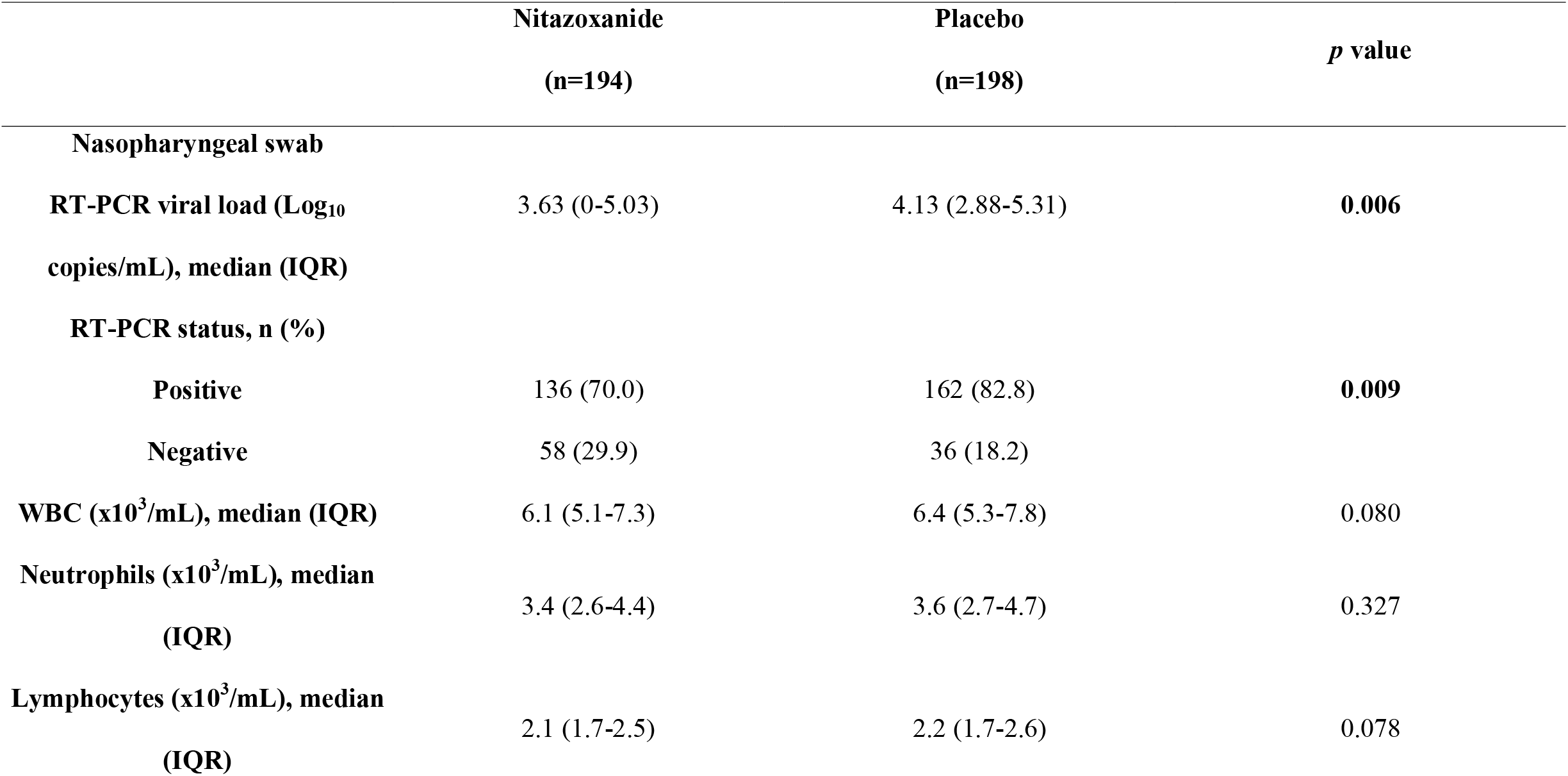
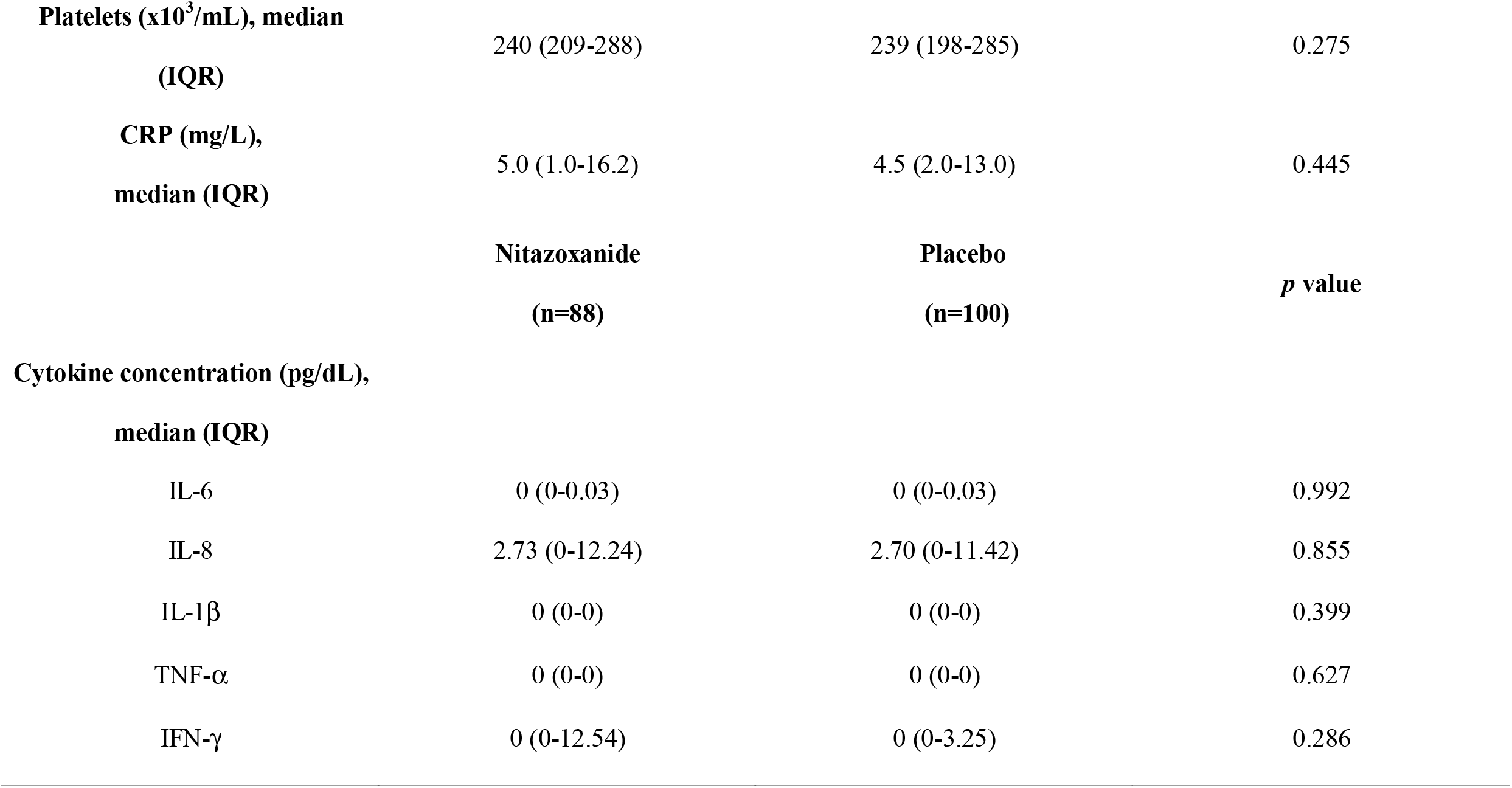
Secondary outcomes after 5 days of therapy.

No deaths or life-threatening adverse events were reported in the nitazoxanide arm. Mild and moderate adverse events were experienced by patients in both arms (nitazoxanide, 30.9%; placebo, 30.4%) during the 5-day course of therapy **(Table 4)**. The most common adverse events (grades 1 and 2) were diarrhea, headache, and nausea, with no significant differences between groups. Four patients reported severe adverse events (headache alone or with diarrhea); however, they were more frequent in the placebo arm. Discolored urine was significantly more frequent in the nitazoxanide arm. Six patients in the nitazoxanide arm and one patient in the placebo arm discontinued therapy due to moderate diarrhea and vomiting **(Table S5)**.

**Table 4.** Adverse Events.

|  | **Nitazoxanide** | **Placebo** | ***p* value** |
| --- | --- | --- | --- |
|  | **(n=194)** | **(n=198)** |  |
| Number of participants with at least one adverse event | 60 (30.9%) | 60 (30.4%) | 0.913 |
| Number of participants with two adverse events | 22 (11.3%) | 18 (9.1%) | 0.507 |
| Number of participants with three or more adverse events | 16 (8.2%) | 12 (6.1%) | 0.438 |
| Number of participants with severe adverse event | 1 (0.5%) | 1 (0.5%) | 0.999 |
| Number of participants discontinuing treatment because of moderate adverse events | 6 (3.1%) | 1 (0.5%) | 0.065 |
| **Detailed adverse events** |  |  |  |
| Diarrhea | 57 (29.4%) | 49 (24.7%) | 0.309 |
| Headache | 34 (17.5%) | 32 (16.1%) | 0.787 |
| Nausea | 28 (14.4%) | 29 (14.6%) | 0.999 |
| Abdominal pain | 10 (5.2%) | 5 (2.5%) | 0.197 |
| Abnormal color of urine | 11 (5.6%) | 3 (1.5%) | 0.031 |
| Vomiting | 9 (4.6%) | 3 (1.5%) | 0.085 |
| Pruritus | 4 (2.1%) | 1 (0.5%) | 0.212 |
| Urticaria | 1 (0.5%) | 3 (1.5%) | 0.623 |

Data are n (%) and include all adverse events reported after nitazoxanide or placebo.

## Discussion

In the current multicenter, double-blind, randomized, placebo-controlled trial in mild Covid-19 patients, we found that symptom resolution (dry cough, fever, and fatigue) did not differ between the nitazoxanide and placebo after 5 days of therapy. However, at the 1-week follow-up phone call, 78% in the nitazoxanide arm and 57% in the placebo arm reported complete resolution of symptoms (*p*=0.048). Nitazoxanide was safe, significantly reduced viral load, and increased the proportion of patients testing negative for SARS-CoV-2 after 5 days of therapy compared to placebo. Nitazoxanide did not prevent hospitalization or effect any change in complete blood count, CRP levels, or serum biomarkers of inflammation.

There is an urgent need for evidence-based pharmacotherapeutics for Covid-19. In the challenging context of a pandemic, drug repurposing can reduce the time-consuming drug development process and allow rapid deployment of effective therapies to the population [2]. Before implementation of this trial, the NIH Clinical Collection (NCC) library, a collection of 727 FDA-approved drugs or drug-like compounds with a history of use in human clinical trials, was screened for potential *in vitro* antiviral activity against SARS-CoV-2 **(See Supplemental Methods and Figure S1)**. Nitazoxanide and tizoxanide, its active metabolite, significantly reduced viral load in VeroE6, human embryonic kidney (HEK 293T), and lung epithelial (Calu-3) cells infected with SARS-CoV-2, without inducing loss of cell viability. Nitazoxanide is a very inexpensive drug and can thus be widely distributed by private and publicly funded health systems [5]. It also has a low production cost, and existing pipelines are readily adaptable to its manufacturing. Its adverse effect profile is well-known, since it has been commercially available and used in the clinic since 1996; indeed, a commercial formulation of nitazoxanide was used in this trial. A dosage regimen of 500 mg every 12 hours is approved and commonly prescribed for treatment of intestinal parasitosis [6], with few reported adverse events. In our study, the same dose was administered, but every 8 hours, based on *in vitro* pharmacological studies by our group and published data on plasma concentration [7-9], in order to maximize potential inhibition of SARS-CoV2 *in vivo* [8]. We assumed that high ratios of maximum serum concentration at doses safe in humans to maximal *in vitro* effective antiviral concentration would translate into higher potential to achieve viral suppression at approved doses [10].

Nitazoxanide failed to meet the primary outcome in patients with mild Covid-19 when evaluated after 5 days of therapy, but when evaluated at the 1-week follow-up, 78% in the nitazoxanide arm and 57% in the placebo arm reported complete resolution of symptoms (*p*=0.048). Consistent with the *in vitro* data, we observed significant reductions in viral load after a 5-day course of nitazoxanide in patients with mild Covid-19. This effect may have epidemiological impact, potentially decreasing community spread of SARS-CoV-2 [11, 12], morbidity [13], and mortality [14].

We enrolled patients with mild Covid-19 as soon as possible after symptom onset because antivirals seem to be more effective at this stage of infection [15]. The cutoff point for enrollment was 3 days after onset of first symptom; however, as therapy was begun only after confirmation of SARS-CoV-2 infection, the median timing of treatment initiation was 5 (4-5) days after symptom onset **(Table 1)**. In previous trials of putative antiviral agents, median time to initiation of treatment was 13 days [16], 9 days [17], 8-9 days [18], and 4-5 days [19] after symptom onset. Initiating therapy even earlier might be more effective, since the peak SARS-CoV-2 viral load usually occurs before symptom onset [20] and systemic hyperinflammation rather than viral pathogenicity dominates later stages of Covid-19, at which point antiviral therapy could be ineffective [20, 21].

Besides timing, two additional points which may have biased our findings in favor of the nitazoxanide arm were the severity of Covid-19 at presentation and the favorable demographic profile of the sample. Only patients with mild Covid-19 were enrolled; most were young adults (aged 18-39), few had comorbidities (12-18%), and use of concomitant medications was infrequent (less than 20% of the sample). Further studies are needed to evaluate whether nitazoxanide may play a role in more advanced Covid-19. In this line, Hung *et al*. recently reported the importance of combining antiviral therapies in patients with severe Covid-19, aiming to reduce viral load and mitigate symptoms [19].

The increase in dosage frequency of nitazoxanide did not result in any significant change in the adverse event rate, although urine discoloration was observed. Neither frequency nor severity of adverse events differed significantly between the arms, suggesting nitazoxanide is a safe therapy for mild Covid-19 patients.

This study has several limitations. A long-term analysis of the effects of therapy (e.g., beyond 28 days) was not performed. Only three symptoms (dry cough, fever, and fatigue) were considered for analysis of primary outcome, even though Covid-19 is now known to have protean manifestations and patients reported other symptoms. Initially, patients were followed during the 5-day course of therapy; only those who continued to have one of the symptoms of interest after completion of therapy were contacted again 1 week later. At this second time point, nitazoxanide therapy was found to have reduced symptom duration significantly as compared to placebo. All patients were instructed to take their study medication 3 times daily as prescribed, return to the study site if symptoms worsened or adverse effects developed, and complete their symptom journals with data on any adverse events, the intensity of each Covid-19 symptom, and when symptoms abated and resolved. However, we cannot ascertain the extent to which patients followed these instructions. Nevertheless, given the placebo-controlled design, nonadherence may have occurred in both groups.

In summary, in patients with mild Covid-19 enrolled within 3 days of symptom onset, nitazoxanide as compared with placebo was not an effective therapy in terms of accelerating symptom resolution after 5 days of therapy. However, at the 1-week follow-up, 78% in the nitazoxanide arm and 57% in the placebo arm reported complete resolution of symptoms (*p*=0.048). Nitazoxanide was safe, significantly decreased viral load, and increased the proportion of patients who tested negative for SARS-CoV-2 after completion of therapy.

## Data Availability

The investigators plan on turning all individual participant data (IPD) available after the publication of the manuscript, with cautious of not sharing confidential data.

## Acknowledgements

The authors thank Moira Elizabeth Schöttler (Rio de Janeiro, Brazil) and Filippe Vasconcellos (São Paulo, Brazil) for editing assistance.

## Contributors

PRMR, PLS, FFC were responsible for the design, analyzing, and writing of the manuscript. MACMJ, PGGMMT, MAM, LFGO, CCL, EZS, WFJ, EM, NFM, JMJG, MNC, ISS, NFP, PVMM were responsible for recruitment and clinical care of the patients. APSMF, KGF, RPR, AFC, PAA, JLM, ATC, DBBT, REM, RRL, JRLS, PP were responsible for the laboratory and statistical analysis. KGF and PP were also responsible for writing of the manuscript. All authors reviewed and approved the final version of the manuscript.

## Conflict of interest

Dr. Rocco reports personal fees from SANOFI as a DSMB member. The other authors declare no competing interests.

## Funding

Supported by the Brazilian Council for Scientific and Technological Development (CNPq), Brazilian Ministry of Science, Technology, and Innovation for Virus Network; Brasília, Brazil, number: 403485/2020-7 and Funding Authority for Studies and Projects, Brasília, Brazil, number: 01.20.0003.00.

## Notes

### Clinical Trial

NCT04552483

### Author Declarations

The IRB/oversight body was the Brazilian National Committee of Ethics in Research (CONEP), and the approval number is CAAE: 32258920.0.1001.5257. The study was further approved by local committees of ethics in research for the seven health units. The investigators have followed all appropriate research reporting guidelines. All necessary patient/participant consent has been obtained and the appropriate institutional forms have been archived.

