## Supplemental for "Early use of nitazoxanide in mild Covid-19 disease: randomized, placebo-controlled trial"

**Table of Contents:**


| | |
|---|---|
| **Committees, Leadership, and Investigators** | **2** |
| **Supplemental *In Vitro* Data to Support Clinical Trial Design** | **5** |
| **Supplemental *In Vitro* Methods** | **5** |
| **Supplemental *In Vitro* Results** | **9** |
| **Supplemental Clinical Methods** | **14** |
| **Additional Details on the Randomization Procedure** | **15** |
| **Additional Details on Data Collection** | **15** |
| **Additional Details on Interventions** | **16** |
| **Additional Details on Outcomes** | **19** |
| **Additional Details on Changes in Protocol During Study** | **20** |
| **Additional Details on Sample Size Calculation** | **20** |
| **Figure S1: Nitazoxanide has antiviral activity against SARS-CoV-2 in cell culture.** | **21** |
| **Figure S2 - Time course of symptoms in patients who tested positive for SARS-CoV-2 treated with nitazoxanide and placebo** | **24** |
| **Table S1- List of Sites and Number of Randomized Patients Per Site** | **25** |
| **Table S2 - Patient Self-Administered Clinical Questionnaire** | **26** |
| **Table S3 - Additional Characteristics of the Population at Baseline** | **30** |
| **Table S4 - Detailed Profile of Hospitalized Patients** | **31** |
| **Table S5 - Adverse Events** | **32** |
| **References** | **33** |

**Committees, Leadership, and Investigators**

**SARITA-2**: The SARITA-2 trial is supported by the Brazilian Ministry of Science, Technology, and Innovation (MCTI), formerly the Ministry of Science, Technology, Innovation, and Communications (MCTIC), and coordinated by the Federal University of Rio de Janeiro (lead investigator: Prof. Patricia R.M. Rocco)

**Executive Committee:** Patricia R. M. Rocco, MD, PhD,[1] Pedro L. Silva, PhD,[1] Fernanda F. Cruz, MD, PhD[1]

**Steering Committee:**
Patricia R. M. Rocco, MD, PhD[1*]; Pedro L. Silva, PhD[1*]; Fernanda F. Cruz, MD, PhD[1*]; Marco Antonio C. M. Junior, MD, MSc[2]; Paulo F. G. M. M. Tierno, MD[3]; Marcos A. Moura, MD, PhD[4]; Luís Frederico G. De Oliveira, MD[5]; Cristiano C. Lima[6]; Ezequiel A. Dos Santos, MSc[7]; Walter F. Junior, BDS[8]; Ana Paula S. M. Fernandes, PhD[9]; Kleber G. Franchini, MD, PhD[10]; Erick Magri[2]; Nara F. de Moraes, MD[3]; José Mário J. Gonçalves[4]; Melanie N. Carbonieri, MD[5]; Ivonise S. Dos Santos, BDS[6]; Natália F. Paes, MSc[7]; Paula V. M. Maciel, MD[8]; Raissa P. Rocha, PhD[9]; Alex F. de Carvalho, PhD[9]; Pedro A. Alves, PhD[11]; José Luiz P. Modena, PhD[12]; Artur T. Cordeiro, PhD[10]; Daniela B. B. Trivella, PhD[10]; Rafael E. Marques, PhD[10]; Ronir R. Luiz, PhD[1]; Paolo Pelosi, MD, FERS,[13, 14] Jose Roberto Lapa e Silva, MD, PhD[1]; on behalf of the SARITA-2 investigators

**Affiliations:** 1. Federal University of Rio de Janeiro, Rio de Janeiro, Brazil 2. Hospital Municipal de Emergências Albert Sabin, São Caetano, São Paulo, Brazil 3. Hospital Municipal de Barueri Dr Francisco Moran, Barueri, São Paulo, Brazil 4. Hospital e Maternidade Therezinha de Jesus, Juiz de Fora, Minas Gerais, Brazil. 5. Hospital Santa Casa de Misericórdia de Sorocaba, Sorocaba, São Paulo, Brazil 6. Secretaria de Estado de Saúde do Distrito Federal, Brasília, Distrito Federal, Brazil 7. Secretaria Municipal de Saúde de Bauru, Bauru, São Paulo, Brazil 8. Secretaria Municipal de Saúde de Guarulhos, Guarulhos, São Paulo, Brazil 9. Centro de Tecnologia de Vacinas, Universidade Federal de Minas Gerais, Belo Horizonte, Minas Gerais, Brazil 10. Brazilian National Biosciences Laboratory (LNBio) and Brazilian Center for Energy and Materials Research (CNPEM), Campinas, Brazil 11. Instituto René Rachou – Fiocruz, Belo Horizonte, Minas Gerais, Brazil, 12. State University of Campinas, Campinas, Brazil. 13. Department of Surgical Sciences and Integrated Diagnostics, University of Genoa, Genoa, Italy, 14. San Martino Policlinico Hospital, IRCCS for Oncology and Neurosciences, Genoa, Italy

**Trial Coordinating Center: Federal University of Rio de Janeiro** (Patricia R. M. Rocco, MD, PhD; Pedro Leme Silva, PhD; Fernanda Ferreira Cruz, MD, PhD).

**Data and Safety Monitoring Board (DSMB):** Prof. Marcus Conde, MD, PhD[1]; Prof. Hugo Caire Faria Neto, MD, PhD[2]; Prof. Hugo Oliveira, MD, PhD.[3]
**Affiliations:**

1. University of Petropolis, Petrópolis, Rio de Janeiro, Brazil
2. FIOCRUZ, Rio de Janeiro, Rio de Janeiro, Brazil
3. Federal University of Rio Grande do Sul, Porto Alegre, Rio Grande do Sul, Brazil

**Site Investigators**
***All trial sites are located in Brazil.***
**Federal University of Rio de Janeiro:** Patricia Rieken Macedo Rocco, Pedro Leme Silva, Fernanda Ferreira Cruz, Cássia Lisboa Braga, Soraia Carvalho Abreu, André Benedito da Silva, Heloísa Leal Setta, Leila Muniz de Almeida Spelta, Ronir Raggio Luiz, José Roberto Lapa e Silva; **Hospital Municipal de Emergências Albert Sabin, São Caetano:** Marco Antonio Cezario de Melo Junior, Erick Magri, Elisabete Ariane da Cunha Mazzola, Edler tertuliano de Almeida lins, , Tatiana Bispo dos Santos, Paula da Silva Pacheco, Matheus Assunção de Assis, Lucineia Mendes Armagni, Francisca Aparecida da Silva Botânico, Amanda Gomes da Silva, Roseli Bernardo Mendes, Marli Monteiro Antonio; **Hospital Municipal de Barueri Dr Francisco Moran, Barueri:** Paulo Fernando Guimarães Morando Marzocchi Tierno, Nara Franzin de Moraes, Simone Luna de Almeida, Ana Paula Silva Araújo, Michelle Cardoso Billet Miranda, Denilda de Jesus Ferreira, Eliana Pereira da Silva, Andreia Avelino Oliveira, Henilton Alves Tonelli, Fernando Carvalho Sacchetin, Igor Silva Fernandes Machado, Ana Lúcia dos Santos Silva; **Hospital e Maternidade Therezinha de Jesus, Juiz de Fora:** Marcos de Assis Moura, Betânia Braga Silva Marques, Patricia Carvalho Pereira Beloti, Ana Luiza Borges Cabral, Leandro Oliveira de Morais, Josias José dos Santos, José Mário de Jesus Gonçalves, Marylin Daniella da Silva, Francisco Lucas Alves Mourão, Karla Letícia Gregório, Rafael de Moura, Marcela Suellen Marques Azalim, Catia Regina Simplício dos Santos, Leonardo Luiz de Almeida Larcher, Dejnane da Cruz Vidal, Letícia da Silva, Heloísa das Mercês Delgado Dultra; **Hospital Santa Casa de Misericórdia de Sorocaba, Sorocaba:** Luis Frederico Gerbase de Oliveira, Fernando Brum Prestes da Silveira, Gabriel Nicolau Bianchi, Fabio Zavarezzi, Melanie Nogueira Carbonieri, Daniele Nunes leite, Ana Paula de Barros Roso, Vania Jaqueline Barbosa, Mariana Mendes, Giliane Aparecida Moraes Silveira Brait, Lysvânia Maria Araújo, Laura Cristina Basílio Oliveira, Aniele Souza Ferreira, Evelyse Bianca Valentim Ramos, Ana Carolina Galati Rodrigues dos Santos; **Secretaria de Estado de Saúde do Distrito Federal, Brasília:** Cristiano Cleidson Lima, Ivonise Sampaio dos Santos, Paulo Ricardo dos Ramos Cardoso, Dina Laine Coutinho de Castro Azevedo, Janaína Pereira Alves, Katherine dos Santos Borges, Steyce Raphaelle Morais Nunes, Zildene dos Santos Moreira Bitencourt, Lívia Batista Silva Carvalho, Marcela Silva Ferreira Fiadeiro, Lissandra Lima, Kelly Cristine Barros Melo, Patrícia Augusto Nunes, Cananda Ferreira Cavalcante, Christiane Silva Pinheiro, Moises Batista de Azevedo, Priscilla Batista Totyda de Menezes, Anna Alice Cardoso Matune, Ana Luiza Laguardia Cantarutti, Mayra Gabrielle Brandão Abrantes, Érica Silene Vernegue, Francisca Maria Pereira, João Francisco de Paula, Bruna de Almeida Silva, Kelly Rodrigues da Costa Silva, Danilo Santana Neiva Gonçalves, Dulcineia Maria dos Reis, Suzaynne Correa Bitencourt Diniz, Alaiene Lelis da Costa Santos, Waldirene Carneiro da Silva, Elaine Angelica Barbosa Elias, Ronaldo Rones Santos da Silva, Leilton Pessoa de Araújo, Waleska Palhares Pires, Damaris Araújo Peixoto, Renata Michele Cassimiro da Silva, Micaela Silva Lopes, Júlia Rodrigues e Rodrigues, Rodrigo Nunes Franco, Vanda de Sousa Nogueira Carvalho, Claudete de Moura Cartaxo, Elianny de Andrade Barros, Eldo Caetano de Moura, Jeanne Alves da Silva, Willian de Araújo Ferreira, Gabriela Alexandra Arrueta Alarcon Canales, Lucilene Alves da Silva, Isabela Amaral Masson, Leandro José Rocha da Silva, Anderson Luiz Akai de Souza, Roseni Saraiva da Silva, Carlos Magno de Olveira da Silva, Mário Roberto Cambraia, Ricardo Stehling, Elisandra da Silva Maciel, Hayane Saraiva de Araújo, Adriane de Oliveira Lemes, Helena Amaral Cotrim Laboissiere, Carolina Mastrodomenico Magdalena Garroti, Angela Rocha de Oliveira Cardoso, Henrique Ferreira dos Santos Junior, Fernando Erick Damasceno Moreira; **Secretaria Municipal de Saúde de Bauru, Bauru:** Ezequiel Aparecido dos

Santos, Natália de Fátima Paes, Carolina Bianchini Trentin Carrer, Flavia Rejane da Silva Carvalho, Natália de Fátima Paes, Fellipe Magela De Araujo, Dayane Paixão Hokama, Daniele Cristina Santarém Sales, Deborah Catherine Salles Bueno, Michele Cristina Vermelho; **Instituto Adolfo Lutz, Bauru:** Virgínia Bodelão Richini Pereira; **Secretaria Municipal de Saúde de Guarulhos, Guarulhos:** Walter Freitas Junior, Paula Veronica Martini Maciel, Silvia Ferreira de Souza, Cleverson Luis Rodrigues, Rodrigo Sarago de Moura, Leandro Jose de Lucas Lima, Amanda Loos Agra Takada, Renart Larry Goda Fernández, Gustavo Jose Le Senechal Salatino, Marta Gomes da Silva, Cleber Alves Carneiro, Marta Lopes Nogueira; **Centro de Tecnologia de Vacinas, Universidade Federal de Minas Gerais:** Ana Paula Fernandes, Alex Fiorini de Carvalho, Raissa Prado Rocha, Leopoldo Ferreira Marques Machado, Vidyleison Neves Camargos, Santuza Maria Ribeiro Teixeira, Flávio Guimarães da Fonseca, Larissa Vuitika, Helena Perez Coelho, Júlia de Souza Reis; **Laboratório de Imunologia de Doenças Virais, Instituto René Rachou – Fiocruz Minas:** Pedro Augusto Alves, Andreza Parreiras Gonçalves e Thaís Bárbara de Souza Silva. **Brazilian National Biosciences Laboratory (LNBio), Brazilian Center for Energy and Materials Research (CNPEM), Campinas:** Rafael Elias Marques, Artur T. Cordeiro, Alexander Borin, Laís D. Coimbra, Gabriela Souza, Stefanie Muraro, Juliana H. C. Smetana, Fabrício F. Naciuk, Matheus Martini, Julia Forato, Leandro O. Bortot, Celso E. Benedetti, Paulo S. Oliveira, Fabiana Granja, Marjorie Bruder, José Luiz Proença Modena, Daniela B. B. Trivella, Kleber G. Franchini. **State University of Campinas, Campinas:** Gabriela Souza, Stefanie Muraro, Matheus Martini, Julia Forato, Fabiana Granja, José Luiz Proença Modena

**Monitoring Service, ATCGen, Campinas:** Carlos Sverdloff, Claudia Viana, Vinicius Rezende, Laura Tavares, Regiane Xavier, Luis Bueno.

**Manufacturer Representatives (Eurofarma):** Cassiano Ricardo de Oliveira Berto, PharmD; Walker Magalhães Lahmann, MD.

**Information Technology Team:** Antonio Jamisson Gonçalves Melo, Rodrigo Ramos Guimarães, José da Silva Junior.

**Supplemental *In Vitro* Data to Support Clinical Trial Design**

**Supplemental *In Vitro* Methods**

Cell lines

Vero CCL81 cells were acquired from the Banco de Células do Rio de Janeiro (BCRJ) repository. HEK293T cells were kindly supplied by Marcio C. Bajgelman (Center for Energy and Materials Research, Campinas, Brazil), and Calu-3 cells were provided by Patricia R.M. Rocco (Federal University of Rio de Janeiro, Brazil). Vero and HEK293T cell lines were cultured in Dulbecco's Modified Essential Medium (DMEM) supplemented with 10% fetal bovine serum (FBS), 1% L-glutamine, and 1% penicillin/streptomycin and maintained at 37 °C in a 5% $CO_2$ atmosphere. Calu-3 cells were cultured in 1:1 DMEM F12 supplemented with 20% FBS, 1% L-glutamine and 1% penicillin/streptomycin, and maintained as described above.

Virus and preparation of viral stock

The SARS-CoV-2 strain HIAE-02 SARS-CoV-2/SP02/human/2020/BRA (GenBank accession number MT126808.1), isolated from a patient diagnosed with Covid-19 in Brazil, was kindly provided by Edison Luiz Durigon (University of São Paulo, Brazil). SARS-CoV-2 stocks were produced by passage in Vero CCL81 cells at 100% confluence using an approximate multiplicity of infection (MOI) of 0.01. Culture supernatant was harvested 36-40 h post-inoculation or on observation of 50% cytopathic effect (CPE), clarified by centrifugation, and stored at −80 °C. Viral stock titer and identity were assessed by viral plaque assay and RT-qPCR.

Compounds

For the *in vitro* assays, the NIH Clinical Collection was obtained from the U.S. National Institutes of Health on a collaborative basis. Stock solutions were maintained at −20 °C, (10mM concentration) in DMSO. Hit quality control was performed by UPLC-MS/MS. For high-throughput screening (HTS) and secondary assays, nitazoxanide was obtained from Sigma-

Aldrich. Tizoxanide (>90% purity by NMR) was obtained by hydrolysis of nitazoxanide using lithium hydroxide (1M) at room temperature, followed by neutralization with hydrochloric acid (1M). The hydrolysis product was filtered, washed, and dried under reduced pressure. Purified compound quality control was performed by melting point measurement, nuclear magnetic resonance ($^{1}H$ and $^{13}C$), and hyphenated ultra-high-performance liquid chromatography-mass spectrometry (UPLC-MS). Compound purity was assessed by NMR using TopSpin 3.6.2 software with trimethylsilyl propionate (TMSP-$d_4$, $D_2O$) as an internal reference, and checked against UPLC-PDA-MS data using the Bruker Data Analysis software.

SARS-CoV-2 cytopathic effect and cell viability assays

Vero CCL81 cells were dispensed in 384-well microplates in suspensions of 1700 cells per well in 45 µL of complete DMEM. Cells were incubated overnight at 37 °C/5% $CO_2$ for adhesion. NIH Clinical Collection stock solutions were used to prepare intermediary plates in complete DMEM at a concentration of 0.05 mM and 2% DMSO before transfer to assay plates. For SARS-CoV-2-induced cytopathic effect assay, 15 µL of compound solutions from intermediary plates were transferred to assay plates containing 45 µL of complete DMEM and cells and infected with SARS-CoV-2 in 15 µL of DMEM at a multiplicity of infection (MOI) of 0.1. For non-infected controls, the previous procedure was applied; however, the cells were mock-infected with 15 µL of DMEM only. Infected and non-infected culture plates were incubated for 60 h at 37 °C, 5% $CO_2$ before staining with Hoechst-33342 2µM and Mitotracker Deep Red 100nM for 45 minutes, following fixation with a 4% PFA solution in PBS.

Imaging and data processing

Plates were imaged with an Operetta automated microscope (Perkin-Elmer). Image segmentation and initial analysis were performed with the Columbus Image Data Storage and Analysis System (Perkin-Elmer). For quantification of the SARS-CoV-2-induced cytopathic effect, one image per well was acquired using the 10× objective lens, and the number of Hoechst-33342 stained nuclei was used to determine the number of cells per well. Normalized inhibition of SARS-CoV-2-induced CPE was calculated by setting the mean values of cells in infected and non-infected control wells as 0 and 100%, respectively.

Antiviral activity assay

The antiviral activity of selected compounds was tested in 24-well plates containing either $2.5\times10^5$ Vero CCL81, $3.5\times10^5$ HEK293T cells, or $2.5\times10^5$ Calu-3 cells. One day (Vero or HEK) or 3 days (Calu-3) after plating, cells were infected with SARS-CoV-2 (MOI 0.01) for 1 h in 5% $CO_2$ at 37 °C. Virus inocula were removed and cell cultures were treated with compounds at 10 μM or 32 μM diluted in complete DMEM, or a serial dilution for concentration-response experiments. Samples were collected at 24 h post-infection (p.i.) in experiments using HEK cells and 48 h when using Vero or Calu-3 cells. Viral load was quantified by RT-qPCR and virus plaque-forming assays.

MTT assay

Cell viability experiments were identical to the antiviral activity assays, except for infection. Cell cultures were grown in 24-well plates, treated with compounds or vehicle, and incubated with the tetrazolium dye MTT (3-(4,5-dimethylthiazol-2-yl)-2,5-diphenyltetrazolium bromide) for 3 hours in 5% $CO_2$ at 37 °C. The supernatant was removed, and tetrazolium crystals were solubilized in DMSO. Absorbance was measured in an EnSpire® Multimode Plate Reader at 490 nm. Cell culture viability data were normalized to vehicle-treated (DMSO) cell culture values and expressed as percentage relative to control.

Virus plaque-forming assay

Supernatant samples were assessed for the presence of infective SARS-CoV-2 viral particles using a plaque assay in Vero CCL81 cells. Confluent cell cultures in 24-well plates were incubated with a 10-fold serial dilution of the sample and plated at 37 °C, 5% $CO_2$, for 1 h. Samples were replaced with a semisolid overlay medium (1% w/v carboxymethylcellulose in DMEM supplemented with 5% FBS) for 3-4 days. The overlay medium was discarded, plates were fixed in 8% w/v paraformaldehyde, and stained with a 1% w/v methylene blue solution. Viral lysis plaques were counted, corrected by the sample dilution factors, and expressed as plaque-forming units (PFU) per mL of supernatant.

Viral RNA extraction and quantification by RT-qPCR

Cell supernatants were collected. Viral RNA extraction was performed using the PureLink RNA Mini Kit (Invitrogen), following manufacturer recommendations, and analyzed with a Nanodrop One spectrophotometer (Thermo Fisher Scientific) before use. SARS-CoV-2 RNA quantification was performed by One-step RT-qPCR according to the Charité protocol[7] using primers and probes for the E gene (forward: 5′-ACA GGT ACG TTA ATA GTT AAT AGC GT-3′, reverse: 5′-ATA TTG CAG CAG TAC GCA TAC GCA CAC A-3′, probe: 5′-6FAM-ACA CTA GCC ATC CTT ACT GCG CTT CG-QSY-3′). All reactions were assembled in a final volume of 12 μL with 3 μL of TaqMan Fast Virus 1-Step Master Mix (Applied Biosystems), 800nM and 400nM of primers and probe, respectively, and 6 μL of 100-fold diluted RNA in ultrapure water. The cycling algorithm used in this study was: 1 cycle at 50 °C for 10 minutes, 1 cycle at 95 °C for 2 minutes, followed by 45 cycles at 95 °C for 5 seconds, and 60 °C for 30 seconds in a QuantStudio3 System

(Applied Biosystems). All applicable measures were taken to prevent cross-contamination of samples, and negative and positive control samples were included in all RT-qPCR plates.

For concentration-response curves, the data were normalized and fitted to the normalized log inhibitor vs. concentration-response curve [Y=100/(1+10^((LogIC50-X)*HillSlope)] in GraphPad Prism v8. Curves were plotted as mean ± SEM triplicate values of each concentration point, constituting one independent experiment. The $EC_{50}$ and $EC_{90}$ values (mean ± SEM) reported in this work were calculated from concentration-response curves from 5 independent experiments.

<u>Statistical analysis</u>

For HTS experiments, the Z factor was calculated as $Z = 1 - 3*(SD_{NI\text{-}controls} + SD_{Inf\text{-}controls})/|<NI_{controls}> - <Inf_{controls}>|$ and Spearman correlation applied for two HTS datasets in the Datawarrior software (openmolecules.org). Datasets from experiments involving viral quantification by RT-qPCR or plaque assays were analyzed using the non-parametric Kruskal-Wallis test coupled to Dunn's multiple comparison test, in which experimental groups were compared to the virus-infected, vehicle-treated control group. Data were expressed as mean + 95% confidence interval (CI). All tests were performed in GraphPad Prism v8.4.0 (GraphPad Software, La Jolla, CA, USA). Significance was established at $P < 0.05$.

#### Supplemental *In Vitro* Results

##### Nitazoxanide and tizoxanide are candidates for Covid-19 drug repurposing

We explored the repurposing potential of existing drugs for anti-SARS-CoV-2 activity and selected nitazoxanide, a broad-acting antiparasitic and antiviral compound[1,2], as a candidate for clinical testing amongst more than 700 compounds (**Figure S1A-B**). Virology studies relevant to clinical translation were performed in preclinical infection systems using human embryonic kidney (HEK293T) (**Figure S1D-F**) and human pulmonary epithelial (Calu-3) cell lines (**Figure S1G-I**). Both nitazoxanide and tizoxanide have specific antiviral activity against SARS-CoV-2 (**Figure S1C-I**) with an appropriate therapeutic index (**Figure S1J-L**). Notably, the concentrations of nitazoxanide and tizoxanide with *in vitro* anti-SARS-CoV-2 activity were within the concentrations attained with therapeutic dose ranges in healthy volunteers[3], implying that they are likely to achieve target concentrations to suppress SARS-CoV-2 under safe dosing conditions[4]. Beyond its well-documented preclinical antiviral activity, additional advantages of nitazoxanide as a repurposing candidate include its favorable oral bioavailability and tolerability in doses well in excess of the usual therapeutic dose range[3–5]. Nitazoxanide is available worldwide, inexpensive to produce and procure, and safe, with a vast body of clinical data accumulated in clinical trials and postmarketing experience, including over 75 million doses with no serious adverse effects reported[6].

**Screening of the NIH Clinical Collection for anti-SARS-CoV-2 active compounds**

To begin exploring the repurposing potential of existing drugs against SARS-CoV-2 infection, we developed a cell-based infection assay in Vero CCL81 cells scaled for image-based high-throughput screening in a 384-well microtiter plate format. The Vero CCL81 cell line derives from African green monkey kidney cells and has been commonly used as an *in vitro* model for viral infections. We used this assay to explore the potential *in vitro* antiviral activity of the NIH Clinical Collection (NCC) library, a collection of 727 FDA-approved drugs or drug-like compounds with a history of use in human clinical trials (**Figure S1A**). Each compound was screened at 10 mM for reduction of the SARS-CoV-2 cytopathic effect (CPE), which is a surrogate readout for viral infection and replication in Vero cells. The primary screen was repeated in two separate replicates (performed on separate days) for assay validation, resulting in a Z-factor > 0.6 and Spearman correlation between independent runs of 0.79 (**Figure S1B**). Compounds were ranked on the basis of reduction of SARS-CoV-2-induced CPE (mean values from both HTS runs). Five compounds reduced CPE by more than 60% and were considered hit candidates, including nitazoxanide (which was selected for follow-up *in vitro* studies) and its active metabolite, tizoxanide.

**Nitazoxanide and tizoxanide reduce SARS-CoV-2 replication *in vitro***

To examine if the anti-cytopathic effect of nitazoxanide reflected a reduction in SARS-CoV-2 replication, we developed a secondary assay in which Vero CCL81 cells were seeded in 24-well plates. Both infected and non-infected cells were treated with DMSO, nitazoxanide, or tizoxanide, added immediately after SARS-CoV-2 adsorption. SARS-CoV-2 replication was assessed by quantifying viral RNA levels (viral load) in cell-culture supernatants at 48 h post-infection using RT-qPCR. Six-point (0.1 to 32 mM) dose-response curves of nitazoxanide and tizoxanide confirmed the inhibition of SARS-CoV-2 replication in Vero CCL81 cells (**Figure S1C**). Curves were fitted to extract the $EC_{50}$ and $EC_{90}$ values. Nitazoxanide and tizoxanide presented comparable $EC_{50}$ (6.1 ± 1.1 μM and 4.9 ± 3.3 μM, respectively) and $EC_{90}$ (18.8 ± 4 and 19.0 ± 4, respectively) values against SARS-CoV-2.

Because our primary purpose was to find compounds that could proceed to clinical trials, nitazoxanide and tizoxanide were selected for further *in vitro* testing in Covid-19-relevant human cell lines. Therefore, we examined the ability of these compounds to inhibit SARS-CoV-2 replication in the human cell lines HEK-293T (embryonic and kidney-derived) and Calu-3 (epithelial and lung-derived). Cell cultures were seeded in 24-well plates, inoculated with SARS-CoV-2, and treated with nitazoxanide and tizoxanide at 10 and 32 mM, or with the vehicle DMSO.

Data were obtained from RT-qPCR and virus plaque-forming assays performed with supernatant samples, which provide information on the abundance of viral RNA and infectious viral titers, respectively. Our results indicated that nitazoxanide and tizoxanide reduced SARS-CoV-2 load in both human cell lines (**Figure S1D-I**).

Experiments performed with these compounds at 10 and 32 mM reduced viral RNA levels by at least 10-fold on average **(Figure S1D)**. The reduction in viral RNA levels in cell cultures treated with nitazoxanide or tizoxanide was accompanied by a more significant reduction in infectious SARS-CoV-2 in supernatant samples, in which nitazoxanide or tizoxanide treatment reduced viral titers by approximately 100-fold at 10 μM or to undetectable levels at 32 μM (**Figure S1E**). Representative SARS-CoV-2 plaque-forming assay images in **Figure S1F** illustrate the decrease in number of SARS-CoV-2 lysis plaques (white dots) in samples from experimental groups treated with nitazoxanide or tizoxanide in comparison to the DMSO (vehicle)-treated group. Briefly, virus lysis plaques are observed in vehicle-treated groups up to a $10^{-4}$ sample dilution factor, while nitazoxanide or tizoxanide treatment causes virus lysis plaques to disappear from samples diluted $10^{-2}$ and forward.

Following the results in the HEK293T cell line, treatment of infected Calu-3 cell cultures with nitazoxanide or tizoxanide at 10 μM reduced RNA levels by 3- to 5-fold, while treatment with 32 μM resulted in an at least 18-fold reduction (**Figure S1G**). The antiviral effect of nitazoxanide and tizoxanide was more pronounced on the infective SARS-CoV-2 load, in which treatment with either compound at 10 μM resulted in a 10- to 20-fold reduction, while treatment at 32 μM reduced viral load by approximately 1000-fold (**Figure S1H**). Representative images of the plaque-forming assays used to determine the infective viral load in experiments using Calu-3 cells illustrate the decrease in the number of viral lysis plaques in groups exposed to nitazoxanide or tizoxanide

(**Figure S1I**). Samples from untreated SARS-CoV-2 infected groups show virus lysis plaques up to the $10^{-4}$ dilution factor, which indicates high viral titers. Samples from groups receiving nitazoxanide or tizoxanide at 32μM show fewer lysis plaques at the lower dilution factor ($10^{-1}$), indicating a reduced viral titer.

Non-infected Vero CCL81, HEK293T, and Calu-3 cell culture plates were prepared in parallel to the antiviral assay plates to assess potential *in vitro* toxicity (**Figure S1J-L**). Cell cultures were incubated with nitazoxanide, tizoxanide, or DMSO for 24h or 48h (depending on the cell type) and an MTT assay was performed to assess cell culture viability. The results showed that nitazoxanide and tizoxanide caused no significant reduction in Vero CCL81 (**Figure S1J**), HEK293T (**Figure S1K**), or Calu-3 (**Figure S1L**) cell viability, even at the highest concentration tested (32 μM), in comparison to the non-treated cells. Thus, nitazoxanide and tizoxanide are not toxic to Vero CCL81, HEK293T, or Calu-3 cells at the tested concentrations.

Altogether, our results show that treatment with nitazoxanide or tizoxanide results in a significant decrease in viral RNA levels and infective viral load in cell culture, consistent with inhibition of viral replication, indicating that nitazoxanide and tizoxanide have antiviral activity against SARS-CoV-2.

**Supplemental Clinical Methods**

Inclusion criteria:

1. Patients with one or more of three selected symptoms of Covid-19 (fever and/or dry cough and/or fatigue) of 1 to 3 days' duration.

2. Age 18 years of age or older.

3. Willingness to take the study therapy.

4. Provision of written informed consent (by patient or a health care surrogate).

Exclusion criteria:

1. Negative result on RT-PCR for SARS-CoV-2 in a nasopharyngeal swab specimen collected at admission.
2. Inability to swallow.
3. History of severe liver disease.
4. Chronic kidney disease requiring renal replacement therapy.
5. Severe heart failure (NYHA class 3 and class 4).
6. Severe chronic obstructive pulmonary disease (COPD) (GOLD 3 and 4).
7. Any cancer in the last 5 years.
8. Any known autoimmune disease.
9. Known allergy to nitazoxanide.
10. Nitazoxanide treatment in the last 30 days.
11. Clinical suspicion of tuberculosis or bacterial pneumonia.

**Additional Details on the Randomization Procedure**

The trial statistician, not involved with patient enrollment or care, obtained a computer-generated randomization list (random.org). Participants were randomized (1:1 ratio) using this list to either the control arm (group B, placebo) or the intervention arm (group A, nitazoxanide). The study treatment (A or B) was revealed to the pharmacist only after patients were registered in the system, ensuring proper concealment of the allocation sequence. The designated pharmacist at each study site was the only person aware of group allocation throughout the trial.

**Additional Details on Data Collection**

A secure website was created by the information technology group (see Committees, Leadership, and Investigators) for data entry, validation, collection, and export. Site investigators, ACTGen monitors, and executive committee members were assigned a secure login and encrypted passwords (128-bit hash). The SARITA-2 system was built on ASP.NET MVC5 with an SQL Server database as the general system; ASP.NET MVC5 as the Web layer; DDD architecture with dependency injection and control inversion as the backend; jQuery with Bootstrap as the frontend; and SQL Server with Entity Framework (Migrations) for data entry and access. The system was hosted in Azure App Service, layer S1 (Microsoft Cloud).

Upon registration of a patient in the system, a unique trial identifier and barcode number (for laboratory tests) were generated and the patient was randomly allocated into group A or B, as mentioned above. Forms within the system were divided into sections that allowed registration of: 1 – demographic data (contact information, patient demographic data, general comments) and upload of informed consent forms; 2 – study day 1 (Baseline): symptoms, vital signs, swab collection data, and results; 3 – Patients who test positive for SARS-CoV-2 (result obtained 1-2 days after RT-PCR, returned to the health facility: clinical data, PCR results, blood test results; 4

– After 5 days of therapy: clinical information, PCR results, blood test results; and 5 – One week after completion of therapy: clinical information, when necessary.

**Additional Details on Interventions**

Timeline

At day 1 (baseline), a nasopharyngeal swab was collected from patients for molecular confirmation of SARS-CoV-2 infection by RT-PCR. Up to 48 hours later, RT-PCR results were displayed. If negative, patients would be excluded from the study. If positive, patients were invited to return to the study site for clinical and laboratory evaluation and initiation of treatment (nitazoxanide 500 mg or placebo, every 8 hours for 5 days, as per group allocation). Patients were given a thermometer and instructed to complete a self-administered questionnaire, which consisted of a list of symptoms and scales on which to record their intensity on each day of therapy (Table S2). Patients were also instructed to return to the study site if any adverse event occurred. One day before completion of therapy, patients received a phone call to remind them to come back to the study site on the next day for final evaluation.

Every included patient was followed according to the following data collection plan:

1. Clinical evaluation (fever, dry cough, fatigue): daily during the course of therapy (via self-administered questionnaire).

2. Nasopharyngeal swab collection for viral load assessment by RT-PCR: at baseline and 1 day after completion of the 5-day course of therapy.

3. Complete blood cell count and C-reactive protein (CRP): immediately before the first dose of study drug and 1 day after completion of the 5-day course of therapy.

4. Serum levels of selected proinflammatory mediators: immediately before the first dose of study drug and 1 day after completion of the 5-day course of therapy.

RNA extraction and real-time polymerase chain reaction (qPCR)

Nasopharyngeal swab samples obtained from each patient were collected in a single tube containing 2 mL of guanidine isothiocyanate transport solution, as previously described.[8] Extraction of the total RNA from collected specimens was performed using the QIAamp Viral RNA Mini Kit (Qiagen, USA), following manufacturer protocols. Quantitation of viral RNA was performed by reverse-transcriptase quantitative real-time polymerase chain reaction (RT-qPCR) following the Berlin (Charité) protocol[7], using the Bio Gene Covid-19 PCR kit (Bioclin, Brazil) per manufacturer instructions. The RT-qPCR reaction was performed in a QuantStudio™ 3 or QuantStudio™ 5 Real-Time PCR System (Thermo Fisher, USA). Human RNase P mRNA was used as internal control and to correct the SARS-CoV-2 viral load in each sample by adjusting viral gene Ct values; to correct the Ct value of SARS-CoV-2 E-gene amplification of each sample, Ct values were normalized using the following equation: (sample SARS-CoV-2 E Ct value × sample RNaseP Ct value / plate mean RNaseP Ct value), as per Duchamp et al.[9] Standard curves were produced by using serial 10-fold dilutions of standard synthetic RNA transcripts of SARS-CoV-2 E gene, ranging from 2 to $2 \times 10^5$ copies/µL (Bioclin, Brazil). Absolute quantification of genomic viral load was performed by comparing sample Ct values to the standard curve. All samples were evaluated centrally at a single site (Centro de Tecnologia de Vacinas, Universidade Federal de Minas Gerais, Brazil).

Self-administered patient questionnaire

Throughout the 5-day course of therapy, patients were instructed to keep a symptom journal recording their body temperature and the presence and intensity (on a scale of 1 to 5) of dry cough, myalgia, sore throat, headache, dyspnea, diarrhea, and other symptoms if present.

Destination of blood for complete blood count and quantitation of CRP

Every study site collected blood from patients after the first positive RT-PCR result and at the end of the 5-day course of therapy. Complete blood count and CRP measurement were performed at the local laboratory of each site.

Destination of serum for quantification of proinflammatory mediators

Serum from patients (2 mL) was collected and stored in a -20 °C freezer at each study site. Cryotubes were labeled with the patient's unique trial identifier and the date of specimen collection. Samples were transported to the biorepository located at the Laboratory of Pulmonary Investigation, Carlos Chagas Filho Biophysics Institute, Federal University of Rio de Janeiro. There, they were stored at -80 °C for molecular analysis of inflammatory mediators.

Molecular analysis

Serum from patients was evaluated for the following biomarkers: interleukin (IL)-6, IL-8, IL-1β, tumor necrosis factor (TNF)-α, and interferon (IFN)-γ. All were measured with commercially available ELISA kits, following the manufacturer's recommendations (Peprotech Inc., Ribeirão Preto, São Paulo, Brazil).

**Additional Details on Outcomes**

This study evaluated efficacy and safety outcomes.

Primary outcomes:

1. Duration (in days) of fever and/or cough and/or fatigue in patients with confirmed Covid-19 treated with nitazoxanide or placebo.

Secondary outcomes:

1 - Evolution of viral load in nasopharyngeal swab specimens in patients with Covid-19 treated with nitazoxanide or placebo at baseline (i.e., at the time of enrollment) and 1 day after completion of the 5-day course of therapy.

2 - Hospitalization rate of patients with Covid-19 treated with nitazoxanide vs. those treated with placebo, over a 14-day period.

3 – Levels of inflammatory mediators (IL-6, IL-1β, IL-8, TNF-α, IFN-γ) in patients with Covid-19 treated with nitazoxanide vs. those treated with placebo, before the first dose of study drug and 1 day after completion of the 5-day course of therapy.

4 – Complete blood count of patients with Covid-19 treated with nitazoxanide vs. those treated with placebo, before the first dose of study drug and 1 day after completion of the 5-day course of therapy

5 - C-reactive protein (CRP) levels of patients with Covid-19 treated with nitazoxanide vs. those treated with placebo, before the first dose of study drug and 1 day after completion of the 5-day course of therapy

Safety outcomes:

1. Incidence of adverse events (AEs) throughout the study.

2. Rate of treatment discontinuation due to AEs.

All outcomes were assessed by blinded investigators. We conducted source data verification of the D8 assessment from study sites and laboratory forms for all patients at sites.

**Additional Details on Changes in Protocol During Study**

Initially, we planned on following patients until D8, regardless of final RT-PCR test result. However, the follow-up period was revised: patients who remained symptomatic 1 day after completion of the 5-day course of therapy were followed for a further week, then contacted by telephone for assessment of clinical criteria and medical advice.

**Additional Details on Sample Size Calculation**

Calculation of the sample size was based on a previous study which demonstrated that 78% of Covid-19 patients in group 4 (Hospitalized without oxygen therapy), according to the WHO ordinal classification, experienced complete resolution of symptoms after receiving placebo.[10] In the present trial, patients were classified as group 2 (Symptomatic and independent), and a greater degree of recovery as measured by symptom-free days (80%) was expected even after placebo. Thus, assuming an 11% increase in symptom-free days in those patients who would receive nitazoxanide compared to placebo, we would need approximately 196 patients per experimental group, admitting a beta error of 15% and alpha error of 5%, for a total n of 392 patients.

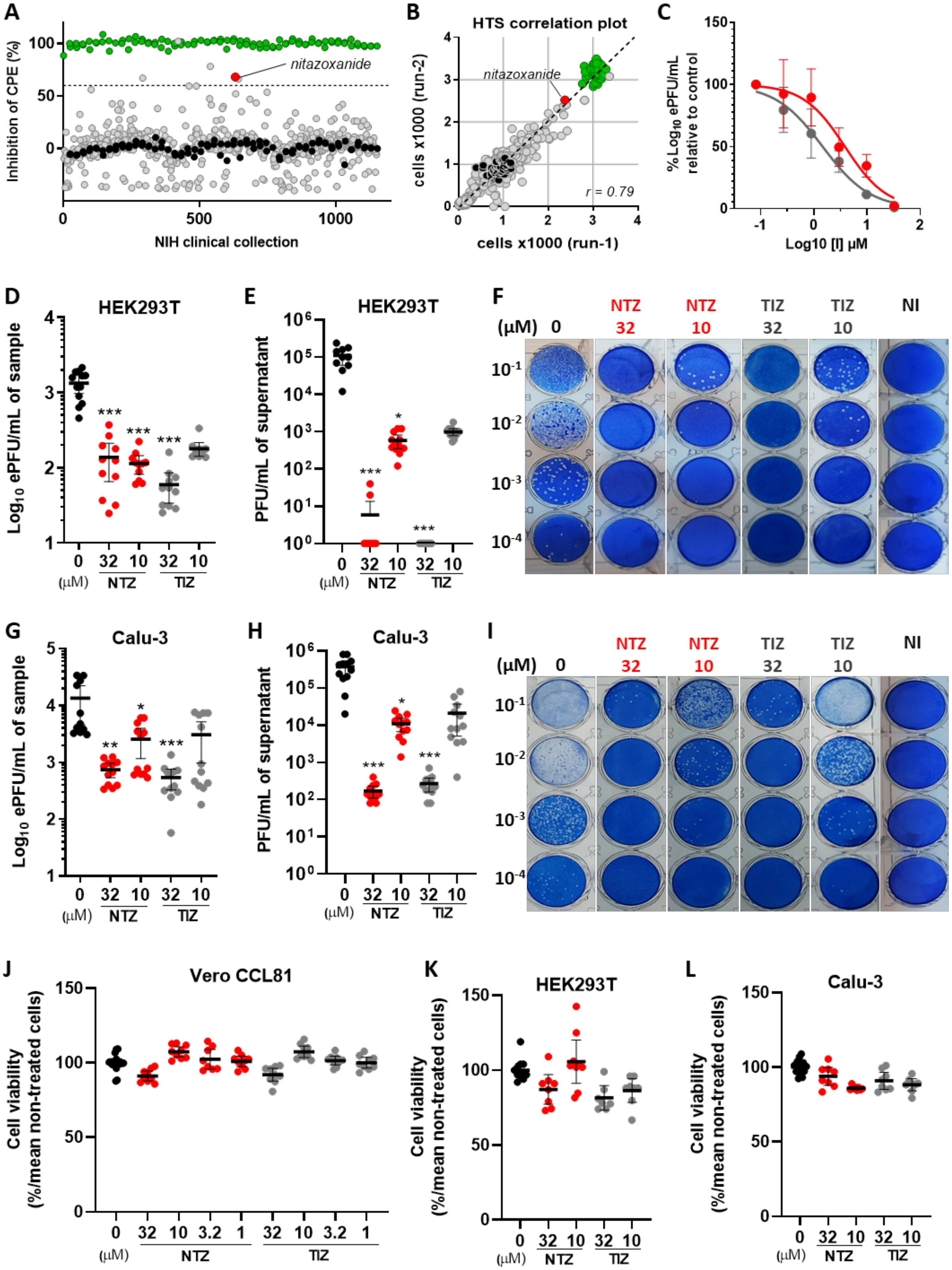
A
Inhibition of CPE (%)
nitazoxanide
NIH clinical collection
B
HTS correlation plot
nitazoxanide
cells x1000 (run-2)
cells x1000 (run-1)
r = 0.79
C
% Log10 ePFU/mL relative to control
Log10 [I] µM
D
HEK293T
Log10 ePFU/mL of sample
***
(µM) NTZ TIZ
E
HEK293T
PFU/mL of supernatant
F
NTZ NTZ TIZ TIZ NI
(µM) 0 32 10 32 10
G
Calu-3
Log10 ePFU/mL of sample
H
Calu-3
PFU/mL of supernatant
I
NTZ NTZ TIZ TIZ NI
(µM) 0 32 10 32 10
J
Vero CCL81
Cell viability (%/mean non-treated cells)
K
HEK293T
L
Calu-3

**Figure S1: Nitazoxanide has antiviral activity against SARS-CoV-2 in cell culture.** **(A)** Screening of the NIH Clinical Collection (NCC) for inhibitors of SARS-CoV-2-mediated cytopathic effect (CPE) in Vero CCL81 cells. The 727 NCC compounds (grey circles) were assayed and compared to non-infected (100%, green) and untreated (0%, black) controls to calculate the percentage of CPE inhibition. Compounds that inhibited CPE >60% were considered hit candidates (nitazoxanide shown in red). **(B)** Correlation plot between two independent HTS experiments. **(C)** Concentration-response curves of nitazoxanide and tizoxanide in SARS-CoV-2 infected Vero CCL81 cells (MOI 0.01). Viral load was assessed in the supernatant by RT-qPCR. **(D)** Viral load measured by RT-qPCR in supernatant samples from infected HEK293T cells treated or not with nitazoxanide or tizoxanide, at 24h post-infection (p.i.) **(E)** Infectious viral load assessed by plaque-forming assay in supernatant samples from infected HEK293T cell cultures. **(F)** Methylene blue-stained wells representative of the plaque-forming assay using HEK293T cell culture samples, which were serially diluted ($10^{-1}$ to $10^{-4}$) to visualize virus lysis plaques (white dots). **(G)** Viral load measured by RT-qPCR in supernatant samples from infected Calu-3 cells treated or not with nitazoxanide or tizoxanide, at 48 h p.i. **(H)** Infectious viral load was assessed by plaque-forming assay in supernatant samples from infected Calu-3 cell cultures. **(I)** Methylene blue-stained wells representative of viral plaque-forming assay in Calu-3 cell culture samples. which were serially diluted ($10^{-1}$ to $10^{-4}$) to visualize virus lysis plaques (white dots). Nitazoxanide and tizoxanide toxicity were assessed in **(J)** Vero CCL81, **(K)** HEK293T, or **(L)** Calu-3 cell cultures using the MTT assay. NI: non-infected; NTZ: nitazoxanide; TIZ: tizoxanide. *$p<0.05$, **$p<0.01$, ***$p<0.001$ relative to the virus-infected control group. Data in graphs are presented as mean + 95% CI. Data in **(C)** are presented as mean ± SEM and are representative of 5 independent experiments (n=9). Data in **(D, E, G, H)** are representative of 2 independent experiments (n=12). Data in **(J-L)** are representative of at least 2 independent experiments (n=8-18).

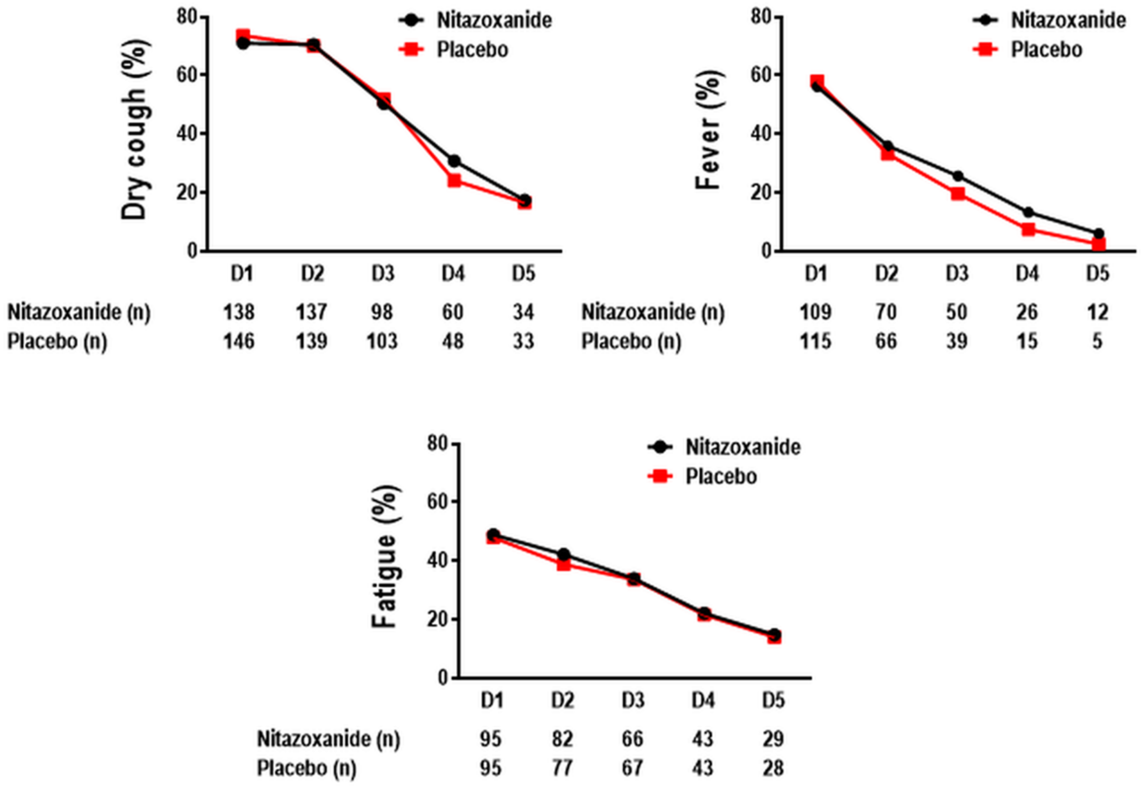


**Figure S2.** Time course of symptoms in patients who tested positive for SARS-CoV-2 treated with nitazoxanide (black line) and placebo (red line). Each symbol represents the percentage of patients with dry cough, fever, and fatigue during 5 days of therapy. n=absolute number of participants with each symptom at each day. Generalized estimating equation (GEE) was used to investigate the effect of time point and group on each variable. No significant differences were observed. Dry cough (p=0.879), fever (p=0.960) and fatigue (p=0.746).

**Table S1. List of Sites and Number of Randomized Patients Per Site**

| | Site | Number of randomized patients |
|---|---|---|
| 1 | Hospital Municipal de Emergências Albert Sabin, São Caetano, São Paulo, Brazil | 116 |
| 2 | Hospital Municipal de Barueri Dr Francisco Moran, Barueri, São Paulo, Brazil | 85 |
| 3 | Hospital e Maternidade Therezinha de Jesus, Juiz de Fora, Minas Gerais, Brazil | 71 |
| 4 | Hospital Santa Casa de Misericórdia de Sorocaba, Sorocaba, São Paulo, Brazil | 68 |
| 5 | Secretaria de Estado de Saúde do Distrito Federal, Brasília, Distrito Federal, Brazil | 33 |
| 6 | Secretaria Municipal de Saúde de Bauru, Bauru, São Paulo, Brazil | 10 |
| 7 | Secretaria Municipal de Saúde de Guarulhos, Guarulhos, São Paulo, Brazil | 9 |

### Table S2 – Patient Self-Administered Clinical Questionnaire

#### DIÁRIO DO PACIENTE

#### #500VoluntariosJa #CombateCOVID19

Para que possamos estar perto de você durante o estudo clínico da NITAZOXANIDA no combate a COVID-19, iremos encaminhar, diariamente, 07 perguntas por SMS para acompanhar seus sintomas durante os próximos 05 dias.

Adicionalmente, pedimos que as mesmas informações que foram enviadas pelo SMS sejam anotadas (com um "x") aqui no seu Diário e que seja entregue no último dia na sua consulta final.

NOME: ______________________________ CPF: _______________

DATA DE CONFIRMAÇÃO POSITIVO PARA COVID-19: ____ / ____ / ____

**USO DO MEDICAMENTO**

**500 mg de NITAZOXANIDA de 8 em 8 horas durante 5 dias consecutivos.**
**ATENÇÃO: Seguir instruções da embalagem do medicamento**

**DATA: ____ / ____ / ____** **D1**

1) Por favor, utilize o termômetro e informe a sua **temperatura** corporal?
( ) Menor que 37º ( ) 37º a 37,5º ( ) 37,6º a 38º ( ) acima de 38º

2) Qual a intensidade da sua **TOSSE SECA**?
Sendo (1) sem tosse seca e (5) tosse de intensidade insuportável.
(1) (2) (3) (4) (5)

3) Qual a intensidade da sua **DOR DE GARGANTA**?
Sendo (1) nenhuma dor e (5) dor de garganta de intensidade insuportável.
(1) (2) (3) (4) (5)

4) Qual a intensidade da sua **DOR DE CABEÇA**?
Sendo (1) nenhuma dor e (5) dor de cabeça de intensidade insuportável.
(1) (2) (3) (4) (5)

5) Qual a intensidade das suas **DORES MUSCULARES**?
Sendo (1) nenhuma dor e (5) dor de intensidade insuportável.
(1) (2) (3) (4) (5)

6) Qual a intensidade da seu **DESCONFORTO RESPIRATORIO**?
Sendo (1) nenhum desconforto e (5) desconforto de intensidade insuportável.
(1) (2) (3) (4) (5)

**DATA: ____ / ____ / ____ D2**

1) Por favor, utilize o termômetro e informe a sua **temperatura** corporal?
( ) Menor que 37º ( ) 37º a 37,5º ( ) 37,6º a 38º ( ) acima de 38º

2) Qual a intensidade da sua **TOSSE SECA**?
Sendo (1) sem tosse seca e (5) tosse de intensidade insuportável.
(1) (2) (3) (4) (5)

3) Qual a intensidade da sua **DOR DE GARGANTA**?
Sendo (1) nenhuma dor e (5) dor de garganta de intensidade insuportável.
(1) (2) (3) (4) (5)

4) Qual a intensidade da sua **DOR DE CABEÇA**?
Sendo (1) nenhuma dor e (5) dor de cabeça de intensidade insuportável.
(1) (2) (3) (4) (5)

5) Qual a intensidade das suas **DORES MUSCULARES**?
Sendo (1) nenhuma dor e (5) dor de intensidade insuportável.
(1) (2) (3) (4) (5)

6) Qual a intensidade da seu **DESCONFORTO RESPIRATORIO**?
Sendo (1) nenhum desconforto e (5) desconforto de intensidade insuportável.
(1) (2) (3) (4) (5)

7) Você apresenta **DIARREIA**?
Sendo (1) nenhum sintoma e (5) diarréia de alta intensidade.
(1) (2) (3) (4) (5)

8)
Sendo (1) nenhum sintoma e (5) diarréia de alta intensidade.
(1) (2) (3) (4) (5)

**DATA: ____ / ____ / ____ D3**

1) Por favor, utilize o termômetro e informe a sua **temperatura** corporal?
( ) Menor que 37º ( ) 37º a 37,5º ( ) 37,6º a 38º ( ) acima de 38º

2) Qual a intensidade da sua **TOSSE SECA**?
Sendo (1) sem tosse seca e (5) tosse de intensidade insuportável.
(1) (2) (3) (4) (5)

3) Qual a intensidade da sua **DOR DE GARGANTA**?
Sendo (1) nenhuma dor e (5) dor de garganta de intensidade insuportável.
(1) (2) (3) (4) (5)

4) Qual a intensidade da sua **DOR DE CABEÇA**?
Sendo (1) nenhuma dor e (5) dor de cabeça de intensidade insuportável.
(1) (2) (3) (4) (5)

5) Qual a intensidade das suas **DORES MUSCULARES**?
Sendo (1) nenhuma dor e (5) dor de intensidade insuportável.
(1) (2) (3) (4) (5)

6) Qual a intensidade da seu **DESCONFORTO RESPIRATORIO**?
Sendo (1) nenhum desconforto e (5) desconforto de intensidade insuportável.
(1) (2) (3) (4) (5)

7) Você apresenta **DIARREIA**?
Sendo (1) nenhum sintoma e (5) diarréia de alta intensidade.
(1) (2) (3) (4) (5)

| **DATA:** ____ / ____ / ____ | **D4** |
|---|---|

1) Por favor, utilize o termômetro e informe a sua **temperatura** corporal?
( ) Menor que 37º ( ) 37º a 37,5º ( ) 37,6º a 38º ( ) acima de 38º

2) Qual a intensidade da sua **TOSSE SECA**?
Sendo (1) sem tosse seca e (5) tosse de intensidade insuportável.
(1) (2) (3) (4) (5)

3) Qual a intensidade da sua **DOR DE GARGANTA**?
Sendo (1) nenhuma dor e (5) dor de garganta de intensidade insuportável.
(1) (2) (3) (4) (5)

4) Qual a intensidade da sua **DOR DE CABEÇA**?
Sendo (1) nenhuma dor e (5) dor de cabeça de intensidade insuportável.
(1) (2) (3) (4) (5)

5) Qual a intensidade das suas **DORES MUSCULARES**?
Sendo (1) nenhuma dor e (5) dor de intensidade insuportável.
(1) (2) (3) (4) (5)

6) Qual a intensidade da seu **DESCONFORTO RESPIRATORIO**?
Sendo (1) nenhum desconforto e (5) desconforto de intensidade insuportável.
(1) (2) (3) (4) (5)

7) Você apresenta **DIARREIA**?
Sendo (1) nenhum sintoma e (5) diarréia de alta intensidade.
(1) (2) (3) (4) (5)

| **DATA:** ____ / ____ / ____ | **D5** |
|---|---|

1) Por favor, utilize o termômetro e informe a sua **temperatura** corporal?

( ) Menor que 37º ( ) 37º a 37,5º ( ) 37,6º a 38º ( ) acima de 38º

2) Qual a intensidade da sua **TOSSE SECA**?
   Sendo (1) sem tosse seca e (5) tosse de intensidade insuportável.
   (1) (2) (3) (4) (5)

3) Qual a intensidade da sua **DOR DE GARGANTA**?
   Sendo (1) nenhuma dor e (5) dor de garganta de intensidade insuportável.
   (1) (2) (3) (4) (5)

4) Qual a intensidade da sua **DOR DE CABEÇA**?
   Sendo (1) nenhuma dor e (5) dor de cabeça de intensidade insuportável.
   (1) (2) (3) (4) (5)

5) Qual a intensidade das suas **DORES MUSCULARES**?
   Sendo (1) nenhuma dor e (5) dor de intensidade insuportável.
   (1) (2) (3) (4) (5)

6) Qual a intensidade da seu **DESCONFORTO RESPIRATORIO**?
   Sendo (1) nenhum desconforto e (5) desconforto de intensidade insuportável.
   (1) (2) (3) (4) (5)

7) Você apresenta **DIARREIA**?
   Sendo (1) nenhum sintoma e (5) diarréia de alta intensidade.
   (1) (2) (3) (4) (5)

**Table S3. Additional Characteristics of the Population at Baseline**

| | Overall (392) | Nitazoxanide (194) | Placebo (198) | *p* value |
|---|---|---|---|---|
| **Age in years, median (IQR)** | 37 (29-45) | 37 (28-45) | 37 (29-45) | 0.772 |
| **Previous use of medications, n (%)** | | | | |
| Steroids | 23 (5.9%) | 12 (6.2%) | 11 (5.6%) | 0.833 |
| NSAIDs | 8 (2.0%) | 5 (2.6%) | 3 (1.5%) | 0.499 |
| Azithromycin | 39 (9.9%) | 19 (9.8%) | 20 (10.1%) | 1.000 |
| Ivermectin | 8 (2.0%) | 5 (2.6%) | 3 (1.5%) | 0.499 |
| **Vital signs, mean (SD)** | | | | |
| Temperature, °C | 36.4 (0.5) | 36.4 (0.6) | 36.4 (0.5) | 0.494 |
| Systolic blood pressure, mmHg | 127.6 (14.8) | 126.5 (14.1) | 128.8 (15.4) | 0.159 |
| Diastolic blood pressure, mmHg | 81.7 (11.5) | 81.9 (11.8) | 81.5 (11.3) | 0.591 |
| Heart rate, bpm | 85.1 (13.0) | 85.9 (12.5) | 84.3 (13.4) | 0.181 |
| Respiratory rate, bpm | 18.4 (1.8) | 18.5 (1.9) | 18.3 (1.7) | 0.509 |

IQR: interquartile range, NSAIDs: non-steroidal anti-inflammatory drugs; SD: standard deviation;

**Table S4. Detailed Profile of Hospitalized Patients**

| | Arm | Hospitalization setting | Day of treatment initiation | Days elapsed between treatment initiation and hospitalization |
|---|---|---|---|---|
| 1 | Nitazoxanide | General ward | 07/16/2020 | 2 days |
| 2 | Nitazoxanide | ICU | 07/23/2020 | 1 day |
| 3 | Nitazoxanide | General ward | 07/05/2020 | 1 day |
| 4 | Nitazoxanide | General ward | 07/11/2020 | 1 day |
| 5 | Nitazoxanide | ICU | 07/24/2020 | 4 days |
| 6 | Placebo | General ward | 07/11/2020 | Never took study drug |
| 7 | Placebo | General ward | 07/11/2020 | 1 day |
| 8 | Placebo | General ward | 07/22/2020 | Never took study drug |
| 9 | Placebo | General ward | 08/06/2020 | 4 days |
| 10 | Placebo | General ward | 08/02/2020 | 5 days |

ICU, intensive care unit.

**Table S5. Adverse Events**

| Adverse event | Total (n=392) | | Group | | | |
|---|---|---|---|---|---|---|
| | | | A = Nitazoxanide (n=194) | | B = Placebo (n=198) | |
| | n | % | n | % | N | % |
| **Headache** | | | | | | |
| None | 326 | 83.2 | 160 | 82.5 | 166 | 83.8 |
| Mild | 50 | 12.8 | 25 | 12.9 | 25 | 12.6 |
| Moderate | 11 | 2.8 | 8 | 4.1 | 3 | 1.5 |
| Severe | 4 | 1.0 | 1 | 0.5 | 3 | 1.5 |
| **Diarrhea** | | | | | | |
| None | 287 | 73.2 | 138 | 71.1 | 149 | 75.3 |
| Mild | 68 | 17.3 | 36 | 18.6 | 32 | 16.2 |
| Moderate | 35 | 8.9 | 19 | 9.8 | 16 | 8.1 |
| Severe | 2 | 0.5 | 1 | 0.5 | 1 | 0.5 |
| **Nausea** | | | | | | |
| None | 335 | 85.5 | 166 | 85.6 | 169 | 85.4 |
| Mild | 41 | 10.5 | 19 | 9.8 | 22 | 11.1 |
| Moderate | 14 | 3.6 | 8 | 4.1 | 6 | 3.0 |
| Severe | 2 | 0.5 | 1 | 0.5 | 1 | 0.5 |
| **Vomiting** | | | | | | |
| None | 381 | 97.2 | 186 | 95.9 | 195 | 98.5 |
| Mild | 9 | 2.3 | 6 | 3.1 | 3 | 1.5 |
| Moderate | 2 | 0.5 | 2 | 1.0 | 0 | 0 |
| Severe | 0 | 0 | 0 | 0 | 0 | 0 |
| **Anorexia** | | | | | | |
| None | 388 | 99.0 | 191 | 98.5 | 197 | 99.5 |
| Mild | 3 | 0.8 | 2 | 1.0 | 1 | 0.5 |
| Moderate | 1 | 0.3 | 1 | 0.5 | 0 | 0 |
| Severe | 0 | 0 | 0 | 0 | 0 | 0 |
| **Pruritus** | | | | | | |
| None | 387 | 98.7 | 190 | 97.9 | 197 | 99.5 |
| Mild | 3 | 0.8 | 3 | 1.5 | 0 | 0 |
| Moderate | 1 | 0.3 | 1 | 0.5 | 0 | 0 |
| Severe | 1 | 0.3 | 0 | 0 | 1 | 0.5 |
| **Urticaria** | | | | | | |
| None | 388 | 99.0 | 193 | 99.5 | 195 | 98.5 |
| Mild | 4 | 1.0 | 1 | 0.5 | 3 | 1.5 |
| Moderate | 0 | 0 | 0 | 0 | 0 | 0 |
| Severe | 0 | 0 | 0 | 0 | 0 | 0 |
